# Epistemic Trust, Mistrust and Credulity Questionnaire (ETMCQ) – Validation of the German version in a representative sample

**DOI:** 10.1101/2025.09.03.25331929

**Authors:** Tobias Nolte, Nicola-Hans Schwarzer, David Riedl, Eileen Lashani, Hanna Kamplig, Chloe Campbell, Elmar Brähler, Cedric Sachser, Jörg M. Fegert, Elisabeth Maria Balint, Bernhard Strauss, Katja Brenk-Franz, P. Read Montague, Peter Fonagy, Stephan Gingelmaier, Astrid Lampe, Hannes Kruse

## Abstract

The construct of epistemic trust has garnered significant attention, both conceptually in relation to societal shifts in trust in communicated knowledge and social learning, and empirically in relation to psychopathology. Recently, the ETMCQ, a self-report tool, was developed to assess individual differences in epistemic stance (ES). This paper reports on the validation of a German version of the ETMCQ. Using a representative sample of 2,519 participants older than 16 years of age, the primary aim was to test the factorial validity of the instrument, while also examining associations with age, gender, and education level. A secondary aim explored associations between the three ETMCQ dimensions (trust, mistrust, and credulity) and retrospectively reported childhood maltreatment and other adversity, as well as other psychological factors, including psychopathology.

Exploratory and confirmatory factor analyses revealed three correlated but distinct factors—Trust, Mistrust, and Credulity—largely in line with the original validation, resulting in a 12-item version for the German adaptation. Our findings support previous theoretical links between epistemic stance and psychological functioning, particularly the association between epistemic disruption (high mistrust and/or credulity) and increased symptomatology. Additionally, both Mistrust and Credulity were linked to childhood maltreatment, attachment avoidance, attachment anxiety, and fearful attachment.

Key results suggest associations between ETMCQ factors and developmental psychopathology constructs, with these factors partially mediating the relationship between early adversity and current mental health symptoms. In terms of discriminant validity, we provide ES cut-offs in relation to widely used psychopathology screening tools. Differences in ES were also observed between individuals from the former East and West Germany, as well as in relation to income, gender, and education, suggesting cultural and socio-economic influences on the construct.

In light of these findings, the ETMCQ can be seen as a brief and easy-to-administer tool that holds promise for enhancing clinical and theoretical understanding of interpersonal knowledge transfer.

## Introduction

The construct of epistemic trust (ET), defined as "the capacity of the individual to consider knowledge conveyed by others as significant, relevant to the self, and generalisable to other contexts" (Fonagy & Allison, 2014), has gained increased attention due to its conceptual link with cultural knowledge transfer and social learning. Beyond its developmental implications, ET has been identified as crucial for understanding an individual’s openness to new interpersonal experiences and learning from others—about both the self and others—in psychosocial interventions (Fonagy et al. 2017). As proposed by Liotti (2023), epistemic stance (ES), characterised by three possible dimensions: trust, mistrust, and credulity, originates from various disciplines, including epistemology (Origgi, 2012), philosophy (Fricker, 2007), and sociology (Beck, Giddens & Lash, 1994). This construct offers insights into individuals’ relational lives from developmental, social, pedagogical, and clinical perspectives.

More recently, it has been suggested that ET and epistemic disruption (i.e., heightened epistemic mistrust and/or credulity) may serve as critical factors influencing the development and maintenance of mental ill-health (Bateman et al., 2023; Fonagy, Luyten & Allison, 2015; Luyten et al., 2020;; Nolte et al., 2023). Preliminary evidence indicates that epistemic disruption is linked to psychopathology (Kampling et al., ; Kumpsaoglu et al., under review; 2022; Wendt et al., 2021) and may predict symptom changes in individuals undergoing treatment for complex psychological needs (Riedl et al., 2023, Riedl et a., 2024).

Drawing on the framework of natural pedagogy (Csibra & Gergely 2005, 2006a, 2006b) and Sperber et al.’s (2010) concept of epistemic vigilance, Fonagy and Allison proposed a developmental model where ET plays a central role in determining how individuals receive, believe, internalise, and utilise information gained through communication. The authors link this process to the individual’s broader environment, particularly attachment relationships, as key in transmitting social and relational knowledge (Bowlby, 1979). In these relationships, being mentalised by another and addressed through ostensive cues (e.g., turn-taking, eye contact, a raised eyebrow, specific vocal modulations) creates a heightened state of attention in the receiver, alerting them to the significance of subsequent knowledge transfer. This fosters shared intentionality, increasing the likelihood of communicative intent being perceived as trustworthy and personally relevant. Consequently, the default state of epistemic vigilance in the receiver is modified, facilitating the (transgenerational) transfer of information (Csibra & Gergely, 2006; Gergely & Király, 2019; Gergely & Watson, 1996).

Recently, Campbell et al. (2021) developed a 15-item instrument, the first self-report measure of aspects of an individual’s epistemic stance. They identified a three-factor solution, demonstrating good reliability and validity for the measure. These findings were corroborated and extended by the evaluation of the Italian version of the ETMCQ, with both studies suggesting that a person’s epistemic stance comprises three dimensions (Campbell et al., 2021; Liotti et al., 2023) and by an initial German validation (Weiland et al., 2024). The first dimension, epistemic trust, refers to the openness to consider and trust information communicated interpersonally. It has been suggested that one advantage of secure attachment is the creation of a template that helps a child discern whom to trust and when (Fonagy et al., 2017b), with both trait and state characteristics allowing for context- and relationship-dependent appraisals of communication.

There is some evidence that securely attached individuals appear to adopt an appropriately agentive epistemic stance (Nolte et al., 2023), being better equipped to resist misinformation while recognising social communications that warrant appropriate epistemic trust—a capacity linked to early-life development (Corriveau et al., 2009). Appropriate epistemic trust enhances one’s ability to adapt to the environment and fully benefit from psychosocial learning (Gingelmaier & Schwarzer, 2023).

Epistemic mistrust, by contrast, involves a tendency to appraise information from others as irrelevant, unreliable, or ill-intentioned, leading to relative impermeability to social learning and resistance to interpersonal influence. Dysfunctional epistemic attitudes, possibly as adaptations to early adversity, may result in a pervasive hypervigilance toward others, compromising flexibility in response to changing social conditions and stressors.

The final dimension, epistemic credulity, refers to a lack of discrimination in evaluating new information, leading to increased vulnerability to misinformation, manipulation, and exploitation (Campbell et al., 2021). Heightened levels of mistrust and/or credulity, conceptualised as epistemic disruption, are thought to increase vulnerability to psychopathology (Fonagy et al., 2022), epistemic isolation, and, when viewed through an intersectional lens, epistemic injustice and epistemic violence (Fricker, 2007; Baraitser, 2023). Epistemic disruption, particularly when associated with childhood adversity (Fonagy et al., 2022; Luyten et al., 2019), can inhibit the generation of "we-mode" experiences (e.g., feeling understood and validated by another, such as a therapist or educator) (Fonagy et al., 2022). In such cases, the other is not perceived as having the capacity to understand the communicator’s mind with benevolent intent (Nolte et al., 2023).

Restoring adequate levels of ET or reversing epistemic disruption has been posited as a potential common factor across effective psychosocial interventions, from educational settings (Gingelmaier, Kirsch & Nolte, 2022) to various forms of psychotherapy (Knapen et al.,2021; Nolte et al., 2023). It may also have broader implications for addressing the current societal crisis of trust (Fonagy, personal communication). Recent research employing the ETMCQ has highlighted the impact of epistemic disruption on the failure to create shared understanding, which underpins socially validated reality. Individual differences in compromised social epistemology have been shown to partially account for conspiracy mentality and specific conspiracy beliefs (Brauner et al., 2023; Tanzer et al. 2021, 2024; Kampling et al.2024;).

Despite these promising initial findings, research on the empirical validity of these constructs and their measurement is still in its early stages (Campbell et al. 2021, Liotti et al., 2023). Further psychometric investigation of the ETMCQ, as well as validation studies, are necessary to determine whether it effectively operationalises the proposed constructs and predicts psychopathology and other phenomena underlying individual differences in social learning. Moreover, the role of cultural, socio-economic, and other vulnerability factors in shaping epistemic predispositions, as conceptualised within the framework of intersectionality, remains underexplored.

### The present study

The overall aim of the current study was to validate the ETMCQ in a representative sample of the German population. We report the psychometric properties of the German version of the ETMCQ and examine its associations with key demographic and socio-economic variables. The factor structure of the ETMCQ was assessed using exploratory factor analysis (EFA) and confirmatory factor analysis (CFA), and its reliability was evaluated in this sample. Based on the original instrument’s validation (Campbell et al., 2021) and recent validated versions in other languages (Italian: Liotti et al., 2023; Persian: Asgarizadeh, 2023; French: Greiner, 2024), we anticipated a three-factor structure, with Trust, Mistrust, and Credulity emerging as correlated yet distinct factors.

To investigate the discriminant and convergent validity of the instrument, we assessed the relationship between aspects of epistemic stance, as measured by the three ETMCQ subscales, and retrospectively reported exposure to adverse childhood experiences, psychopathology, attachment, and personality functioning. We tested the following hypotheses:

1. Higher levels of Trust would be negatively associated with childhood adversity, impaired personality functioning, insecure attachment, and psychopathology symptoms, including post-traumatic stress.
2. Both Mistrust and Credulity would be associated with higher levels of childhood adversity, impaired personality functioning, mental health symptoms (including post-traumatic stress), and insecure attachment styles.
3. Given our hypothesis regarding the relationship between epistemic stance, developmental experiences, and psychopathology, we predicted that the three ETMCQ factors would mediate the link between childhood adversity and common psychopathology (e.g., mood disorder symptomatology).

For additional utility, we aimed to establish clinical cut-offs for the three facets of epistemic stance in relation to two commonly used mental health screening instruments (anxiety: GAD-7 and depression: PHQ-9).

## Methods

### Development of the German Epistemic Trust, Mistrust and Credulity Questionnaire

The ETMCQ was first translated into German by three of the authors, with minimal consensus adjustments made where differences arose. An independent translator then back-translated it into English. The accuracy of the initial translation was verified through a qualitative comparison of both versions, and no significant discrepancies were identified.

### Sample and Setting

In collaboration with the independent demographic research institute USUMA Berlin, data were collected from a representative population sample via interviews and self-report questionnaires. Between December 202 and March 2021, 5,418 households across 258 predefined regions were selected using a random route procedure. In households with multiple individuals, one person was randomly selected using the Kish Selection Grid. Survey inclusion criteria were: sufficient German language skills, age above 16, and verbal informed consent. The survey adhered to the Declaration of Helsinki and followed the ethical guidelines of the International Code of Marketing and Social Research Practice of the International Chamber of Commerce and the European Society for Opinion and Marketing Research. Ethical approval was granted by the Ethics Committee of the Medical Faculty of the University of Leipzig (approval number: 474/20-ek).

### Sample Characteristics

A total of 2,519 participants (n = 1,322 female) took part, with ages ranging from 16 to 96 years (M = 50.33, SD = 18.06). For a detailed overview of all sociodemographic data, see Table 1.

**Table 1.**
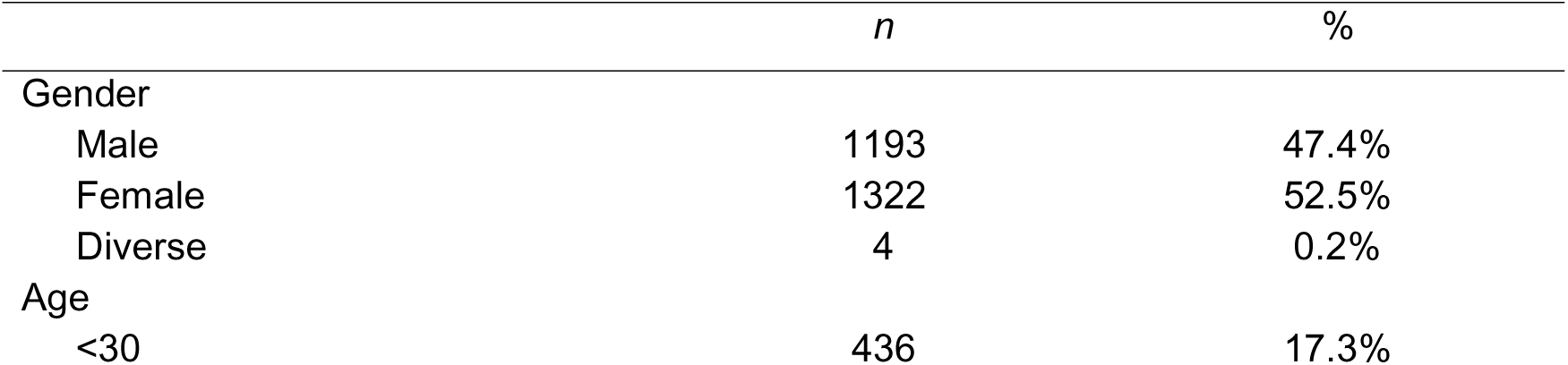
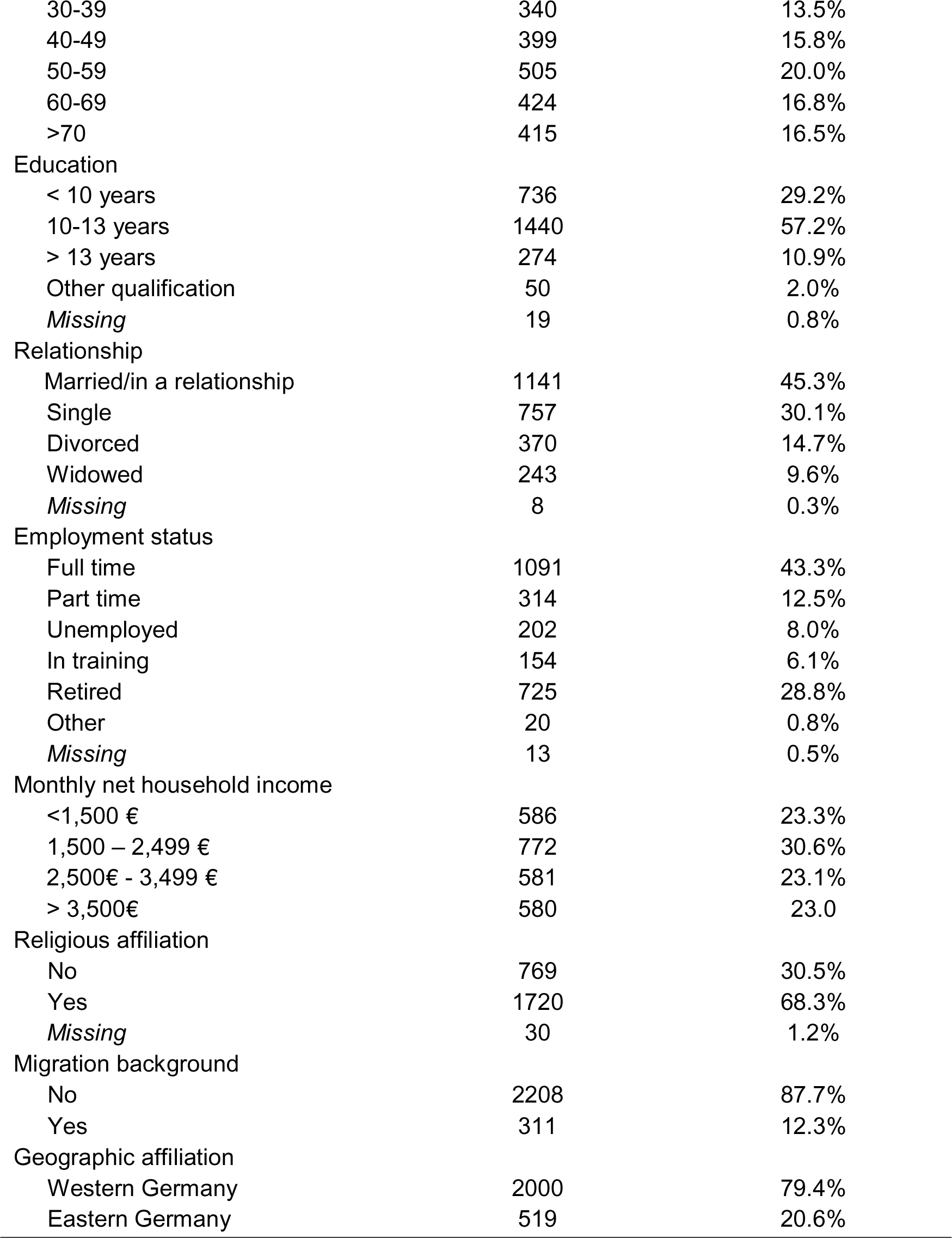
Sociodemographic sample characteristics (*n* = 2519)

|  | *n* | % |
| --- | --- | --- |
| Gender |  |  |
| Male | 1193 | 47.4% |
| Female | 1322 | 52.5% |
| Diverse | 4 | 0.2% |
| Age |  |  |
| <30 | 436 | 17.3% |
| 30-39 | 340 | 13.5% |
| 40-49 | 399 | 15.8% |
| 50-59 | 505 | 20.0% |
| 60-69 | 424 | 16.8% |
| >70 | 415 | 16.5% |
| Education |  |  |
| < 10 years | 736 | 29.2% |
| 10-13 years | 1440 | 57.2% |
| > 13 years | 274 | 10.9% |
| Other qualification | 50 | 2.0% |
| *Missing* | 19 | 0.8% |
| Relationship |  |  |
| Married/in a relationship | 1141 | 45.3% |
| Single | 757 | 30.1% |
| Divorced | 370 | 14.7% |
| Widowed | 243 | 9.6% |
| *Missing* | 8 | 0.3% |
| Employment status |  |  |
| Full time | 1091 | 43.3% |
| Part time | 314 | 12.5% |
| Unemployed | 202 | 8.0% |
| In training | 154 | 6.1% |
| Retired | 725 | 28.8% |
| Other | 20 | 0.8% |
| *Missing* | 13 | 0.5% |
| Monthly net household income |  |  |
| <1,500 € | 586 | 23.3% |
| 1,500 – 2,499 € | 772 | 30.6% |
| 2,500€ - 3,499 € | 581 | 23.1% |
| > 3,500€ | 580 | 23.0% |
| Religious affiliation |  |  |
| No | 769 | 30.5% |
| Yes | 1720 | 68.3% |
| *Missing* | 30 | 1.2% |
| Migration background |  |  |
| No | 2208 | 87.7% |
| Yes | 311 | 12.3% |
| Geographic affiliation |  |  |
| Western Germany | 2000 | 79.4% |
| Eastern Germany | 519 | 20.6% |

### Instruments

#### Adverse Childhood Experiences Questionnaire (ACE)

The ACE (Wingenfeld et al., 2011; Witt et al., 2019) is a widely used self-report instrument for the retrospective assessment of childhood adversities. It consists of ten items, with responses coded as ‘yes’ (1) or ‘no’ (0), covering areas such as emotional, physical, and sexual abuse, emotional and physical neglect, parental separation, witnessing violence against a parent, and familial challenges like substance abuse, mental illness, and incarceration. The cumulative score ranges from 0 to 10. In our sample, the ACE showed high internal consistency (α = 0.81).

#### International Trauma Questionnaire (ITQ)

The ITQ (Cloitre et al., 2018) is a self-report measure that assesses post-traumatic stress disorder (PTSD) and complex PTSD symptoms. The PTSD scale comprises six items assessing three core PTSD dimensions (re-experiencing, avoidance, and heightened perception of threat), along with a three-item scale evaluating functional impairment. For complex PTSD, an additional six items assess disturbances in self-organisation (DSO) across affective dysregulation, negative self-concept, and problematic relationships. All items are rated on a five-point Likert scale from ‘not at all’ to ‘very much’. In our sample, good internal consistency was found for the PTSD (α = 0.89), and DSO (α = 0.87) subscales.

#### Psychological Symptomatology and Distress (PHQ-9, GAD-7, PSS-4)

The PHQ-9 (Löwe et al., 2002; Kliem et al., 2024), the depression module of the Patient Health Questionnaire, assesses the presence and severity of depressive symptoms based on the nine DSM-IV criteria. It is widely used as a screening tool for depression and has demonstrated strong clinical utility (Kocalevent et al., 2013). In our sample, internal consistency for the PHQ-9 was excellent (α = 0.90). The Generalised Anxiety Disorder Scale (GAD-7) (Spitzer et al., 2006; Löwe et al., 2008) is a self-report measure consisting of seven items designed to assess generalised anxiety disorder symptoms according to DSM-IV criteria. The GAD-7 has demonstrated good psychometric properties and clinical usability in German populations (Hinz et al., 2017). In our sample, internal consistency was excellent (α = 0.90).

The Perceived Stress Scale (PSS-4) (Warttig et al., 2013) is a brief screening instrument for evaluating an individual’s perception of stress over the past month. In our sample, internal consistency of the four items was inadequate (α = 0.56), so results should be interpreted tentatively.

#### Operationalised Psychodynamic Diagnosis Structured Questionnaire – 12 Item Version (OPD-SQS)

The OPD-SQS (Ehrenthal et al., 2015) is a 12-item self-report tool designed to assess personality functioning, generating a total score (0–48) and subscale scores (self-perception, interpersonal contact, relationship model; 0–16). Higher scores indicate more severe impairments in personality functioning. The OPD-SQS has demonstrated good validity and reliability (Ehrenthal et al., 2023) and has been linked to future affective well-being and psychosocial impairment (Kerber et al., 2024). In our sample, internal consistency was good to excellent for the total score (α = 0.91) and subscales (α = 0.81–0.87).

#### Experiences in Close Relationships – Revised (ECR-R)

The ECR-R (Ehrenthal et al., 2008) is a 36-item self-report instrument that assesses attachment styles in adult romantic relationships. Participants rate their agreement with each item on a seven-point Likert scale (1 = "strongly disagree" to 7 = "strongly agree"). The instrument measures two key dimensions: attachment-related anxiety (fear of abandonment, intense concern about relationships) and attachment-related avoidance (discomfort with emotional closeness, difficulty trusting others). In our sample, these two dimensions were assessed with one core item each: “I’m afraid that I will lose my partner’s love” for the anxiety scale, and “I prefer not to be too close to romantic partners” for the avoidance scale.

##### Adult Attachment (Self) Rating (AAR)

The Adult Attachment Rating (AAR) is a 21-item self-report measure developed to assess attachment styles in adulthood using prototypical attachment dimensions. This instrument is a brief adaptation of the self-report part of the Adult Attachment Prototype Rating (AAPR; Pilkonis et al., 1994; Strauß & Lobo-Drost, 1999) based upon an item-response analysis (Pilkonis et al., 2014). Participants evaluate each item on a five-point Likert scale, ranging from 0 ("not at all") to 4 ("very much"), to reflect their alignment with various attachment prototypes.

The AAR encapsulates seven subscales derived from the original AAPR, spanning three primary attachment categories: anxious-ambivalent attachment, avoidant attachment, and secure attachment. Specifically, the anxious-ambivalent category is represented by three subscales: Excessive Dependency (characterized by reliance on others for validation and support), Interpersonal Ambivalence (noting fluctuating emotional responses within relationships), and Compulsive Care-Giving (a tendency to adopt caregiving roles to sustain relational closeness). Avoidant attachment is measured through the subscales Rigid Self-Control (favoring emotional self-sufficiency), Defensive Separation (maintaining autonomy through relationship avoidance), and Emotional Detachment (exhibiting limited emotional responsiveness to others’ needs). The Secure Attachment subscale reflects comfort with both intimacy and independence.

Research on the AAPR has established moderate to high reliability across subscales and the total measure, with Cronbach’s alpha values commonly observed between α = .75 and α = .90 (Pilkonis et al., 1994; Simpson & Rholes, 2002). The AAR has shown to reveal convergent validity related to the ECR (self report and informant version and the Adult Attachment Q-Sort). In our sample, reliability was acceptable for the Excessive Dependency (α = .78), Interpersonal Ambivalence (α = .78), and Emotional Detachment (α = .70) subscales. Cronbach’s alpha was questionable for the secure attachment scale (α = .68), and inadequate for Compulsive Care-Giving (α = .52) as well as Defensive Separation (α = .55). For Rigid Self-Control (α = .27), it was insufficient. As the subscales only comprise three items each, we will report results for all of them but warrant cautious interpretation for those with low internal consistency. The AAR was developed as a simple alternative to more elaborate procedures (Pilkonis et al., 2014). Integrating the AAR into this study allows for a streamlined yet robust assessment of adult attachment patterns, providing practical utility in settings where survey administration time is constrained.

### Statistical Analysis

To investigate the factor structure of the ETMCQ, exploratory factor analysis (EFA) was initially performed, followed by confirmatory factor analysis (CFA), as recommended by Campbell et al. (2021). For the EFA, varimax rotation was applied using a maximum likelihood estimator. The number of factors was determined using the scree plot, the elbow criterion, the Kaiser criterion, and the eigenvalue rule (eigenvalues greater than 1). Additionally, a parallel analysis (O’Connor, 2000) was conducted (n = 1,000 parallel datasets, 95th percentile) to confirm the results. CFA, using the maximum likelihood method, was then performed to evaluate a series of models with the following fit indices: χ²-statistic, Root Mean Square Error of Approximation (RMSEA), Standardized Root Mean Square Residual (SRMR), Comparative Fit Index (CFI), and Tucker-Lewis Index (TLI). The following thresholds were used to assess model fit: excellent fit (non-significant χ²-statistic, RMSEA ≤ 0.06, SRMR ≤ 0.06, CFI ≥ 0.95, TLI ≥ 0.95); acceptable fit (non-significant χ²-statistic, RMSEA ≤ 0.08, SRMR ≤ 0.08, CFI ≥ 0.90, TLI ≥ 0.90). Given the large sample size (> 2,500), a significant χ²-statistic was expected. Additionally, the Akaike Information Criterion (AIC) and Schwarz’s Bayesian Information Criterion (BIC) were estimated. The internal consistency of the final factors was evaluated using Cronbach’s alpha and McDonald’s omega coefficients.

All additional statistical analyses were performed using R 4.1.3 (R Core Team, 2022) and SPSS AMOS. Analyses were conducted using all available data, including only participants with complete responses on the relevant variables for a given model. Correlation analyses and group comparisons (t-tests and MANOVAs) were used to explore the relationships between epistemic stance and demographic, as well as psychological, variables. When appropriate, non-parametric alternatives were employed. To test whether the effects of childhood adversity (ACE) on mental health outcomes were mediated by epistemic stance (ETMCQ), mediation models were specified for four mental health outcomes: depression (PHQ-9), anxiety (GAD-7) and perceived stress (PSS-4). Model estimates were computed using a bootstrap procedure (Preacher & Hayes, 2008) with 1,000 resamples.

To establish clinical cut-offs, a Receiver Operating Characteristic (ROC) curve analysis was conducted using scores from the three ETMCQ subscales as predictors for depression and anxiety (based on PHQ-9 and GAD-7 clinical cut-offs). This yielded area under the curve (AUC) values and optimal cut-off scores, balancing specificity (true negative rate) and sensitivity (true positive rate).

## Results

### ETMCQ Factor Structure

In performing an exploratory factor analysis (EFA) on the 15-item version of the ETMCQ, the Kaiser-Meyer-Olkin Measure of Sampling Adequacy (KMO = 0.830) and Bartlett’s Test of Sphericity (χ²(105) = 12,119.72; *p* < .001) indicated that the data were suitable for EFA (Backhaus et al., 2016). The rotated factor loadings suggested a three-factor solution (see Figure 1), accounting for 54.40% of the total variance, consistent with findings from Campbell et al. (2021). Given that the acceptable threshold for factor loadings is typically set at 0.50 (Tabachnick & Fidell, 2012), items ET_3, ET_6, and ET_11 did not meet this criterion and were thus excluded, resulting in a 12-item version. Additionally, high cross-loadings between the Mistrust and Credulity factors for several items suggested the need to evaluate a two-factor model.

**Figure 1.**
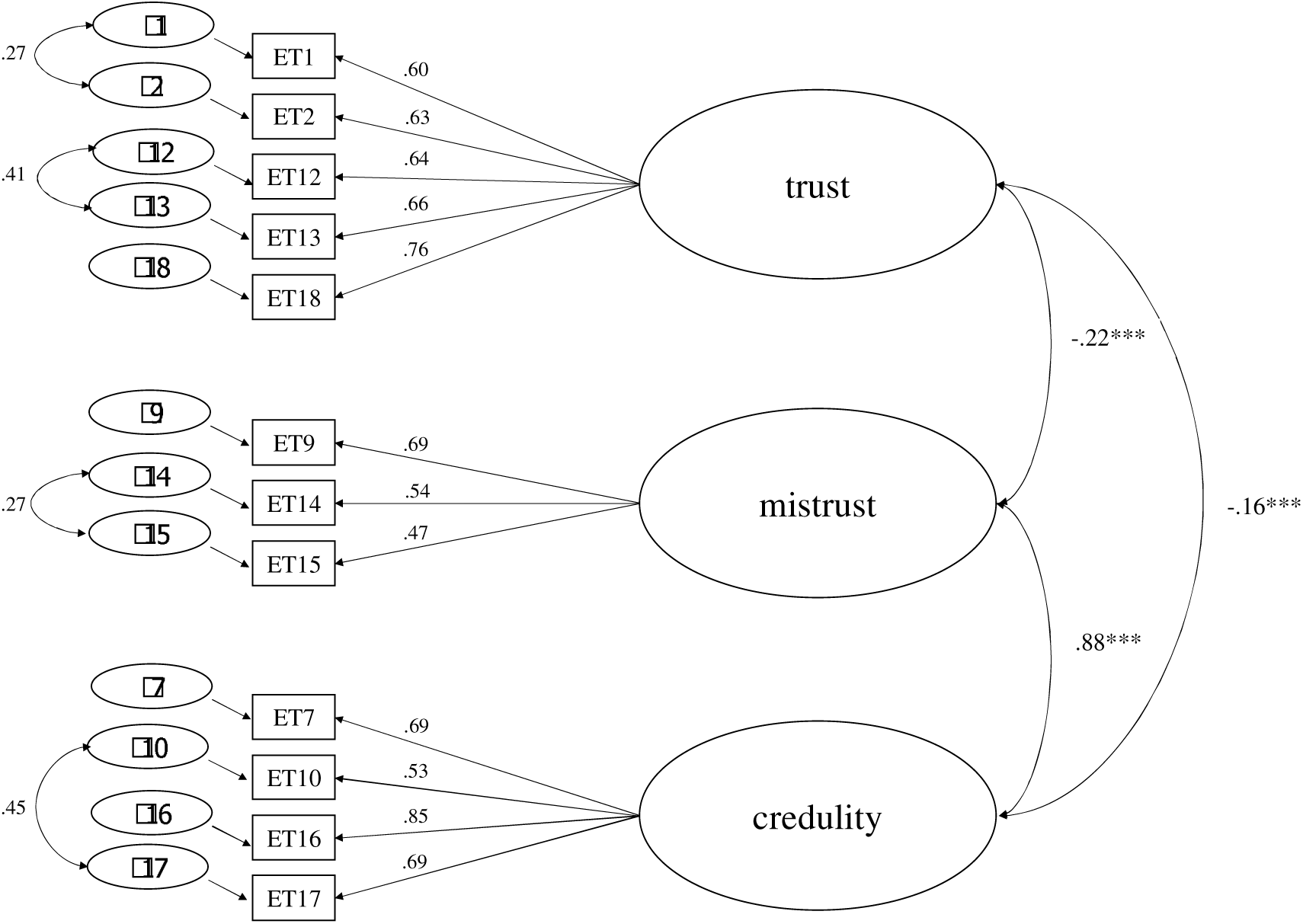
Exploratory factor analysis.

***please add table 2***

### Confirmatory Factor Analysis

The initial model (3 factors, 15 items) proposed by Campbell et al. (2021) was first evaluated (model 1), but the model did not demonstrate good fit to the data (see Table 2). Even after removing two of the lowest-loading items identified by the EFA (ET_3, ET_11), the fit remained inadequate for the 3-factor, 13-item model (model 2). However, upon adding correlated error terms as suggested by Campbell et al. (2021) (ET_1 & ET_2, ET_12 & ET_13, ET_14 & ET_15, ET_10 & ET_17), the fit indices improved to within acceptable ranges (model 3), as shown in Table 2.

We further tested a 3-factor model with the 12 strongest-loading items (model 4), but this model also did not fit the data well. After correlating residuals between several items (model 5), again following Campbell et al. (2021), the model demonstrated good fit (see Table 2). Given the EFA results suggesting a potential two-factor model, we also specified a 2-factor model using the 12 strongest-loading items (model 6), but this model did not fit the data well either. After adding correlated error terms, the fit reached acceptable levels (model 6; see Table 2).

Overall, model 5 (3 factors, 12 items, with correlated error terms) provided the best fit to the data, confirming the discriminant validity as proposed by Campbell et al. (2021). A bi-factorial model was also tested but showed poorer fit indices compared to the best model and was subsequently discarded. We found strong correlations between the Mistrust and Credulity factors (r = 0.57; *p* < .001), while correlations between the Trust and Credulity factors (r = -0.14; *p* < .001) and between the Trust and Mistrust factors (r = -0.16; *p* < .001) were small.

### Internal Consistency

Cronbach’s alpha and McDonald’s omega for the 12-item version of the ETMCQ indicated acceptable reliability for the subscales, given their lengths: the 5-item Trust factor (α = 0.81; C = 0.81), the 3-item Mistrust factor (α = 0.65; C = 0.66), and the 4- item Credulity factor (α = 0.81; C = 0.81). The Mistrust factor was slightly below the conventional 0.70 threshold for acceptability but was considered sufficient due to the short scale length.

**Table 1.** Results of the exploratory factor analysis (rotated) (15 items from original set of 18 items)

|  | Factor loadings |  |  |
| --- | --- | --- | --- |
|  | Mistrust | Trust | Credulity |
| ET_1 | -.158 | .582 | .242 |
| ET_2 | -.161 | .600 | .047 |
| ET_3 | .060 | -.015 | -.218 |
| ET_6 | .397 | -.351 | -.007 |
| ET_7 | .671 | -.032 | .003 |
| ET_9 | .628 | -.106 | .049 |
| ET_10 | .543 | -.051 | .559 |
| ET_11 | .144 | .384 | .273 |
| ET_12 | -.017 | .757 | -.092 |
| ET_13 | -.073 | .768 | -.174 |
| ET_14 | .594 | -.004 | -.178 |
| ET_15 | .522 | -.091 | -.229 |
| ET_16 | .763 | -.065 | -.125 |
| ET_17 | .691 | -.015 | .462 |
| ET_18 | -.030 | .689 | .037 |

The following items were excluded from the final 12-item solution: 3, 6, and 11 ("If someone can show me that something I thought was wrong, I usually change my mind."; "I find it hard to trust information from others with whom I have little in common."; "When I feel good, I am more likely to believe information that someone else has given me." – see Supplements).

**Table 3.** Results of the confirmatory factor analyses.

| Model | $\chi^2$ | CFI | TLI | RMSEA | SRMR | AIC | BIC |
| --- | --- | --- | --- | --- | --- | --- | --- |
| 1. 3-factor, 15 items | $\chi^2(2, n = 2519) = 2459.979, p < 0.001$ | .803 | .762 | .104<br>(.101 – .108) | .0875 | 2525.979 | 2718.422 |
| 2. 3-factor, 13 items | $\chi^2(2, n = 2519) = 1789.491, p < 0.001$ | .847 | .808 | .105<br>(.101 – .109) | .0701 | 1847.491 | 2016.608 |
| 3. 3-factor, 13 items; correlated error terms | $\chi^2(58, n = 2519) = 757.216, p < 0.001$ | .938 | .917 | .069<br>(.065 – .074) | .0571 | 823.216 | 1015.659 |
| 4. 3-factor, 12 items | $\chi^2(51, n = 2519) = 1512.949, p < 0.001$ | .862 | .822 | .107<br>(.102 – .111) | .0580 | 1566.949 | 1724.403 |
| 5. 3-factor, 12 items, correlated error terms | $\chi^2(47, n = 2519) = 478.530, p < 0.001$ | .959 | .943 | .060<br>(.056 – .065) | .0390 | 540.530 | 721.310 |
| 6. 2 factor, 12 items | $\chi^2(53, n = 2519) = 1673.117, p < 0.001$ | .847 | .810 | .110<br>(.106 – .115) | .0622 | 1723.117 | 1868.907 |
| 7. 2 factor, 12 items, correlated error terms | $\chi^2(49, n = 2519) = 524.647, p < 0.001$ | .955 | .940 | .062<br>(.057 – .067) | .0417 | 582.647 | 751.764 |

### Relationship with Demographic Characteristics

When examining the relationship between age and epistemic stance, we found the following correlations: Trust (r = -0.02 [-0.06; 0.02], t(2,494) = -0.01, *p* = 0.360); Mistrust (r = 0.06 [0.03; 0.10], t(2,482) = 3.22, *p* = 0.001); and Credulity (r = 0.00 [-0.03; 0.04], t(2,491) = 0.22, *p* = 0.828).

A MANOVA comparing male and female participants revealed significantly higher Trust and Credulity in women, whereas no significant differences were found for Mistrust (for all associations with demographic characteristics, see Table 2).

Additionally, a higher educational degree was associated with significantly lower epistemic disruption, although no differences were detected for epistemic trust between individuals with and without higher education. Unemployment was linked to reduced Trust and increased epistemic disruption. Similarly, lower household income correlated positively with higher levels of epistemic disruption, and Trust was also lower among those with lower income.

Living in the eastern part of Germany (former German Democratic Republic) was associated with higher levels of Trust, higher levels of Mistrust, and lower levels of Credulity. (M)ANOVA results for sociodemographic variables and epistemic stance are presented in Table 4.

**Table 4.**
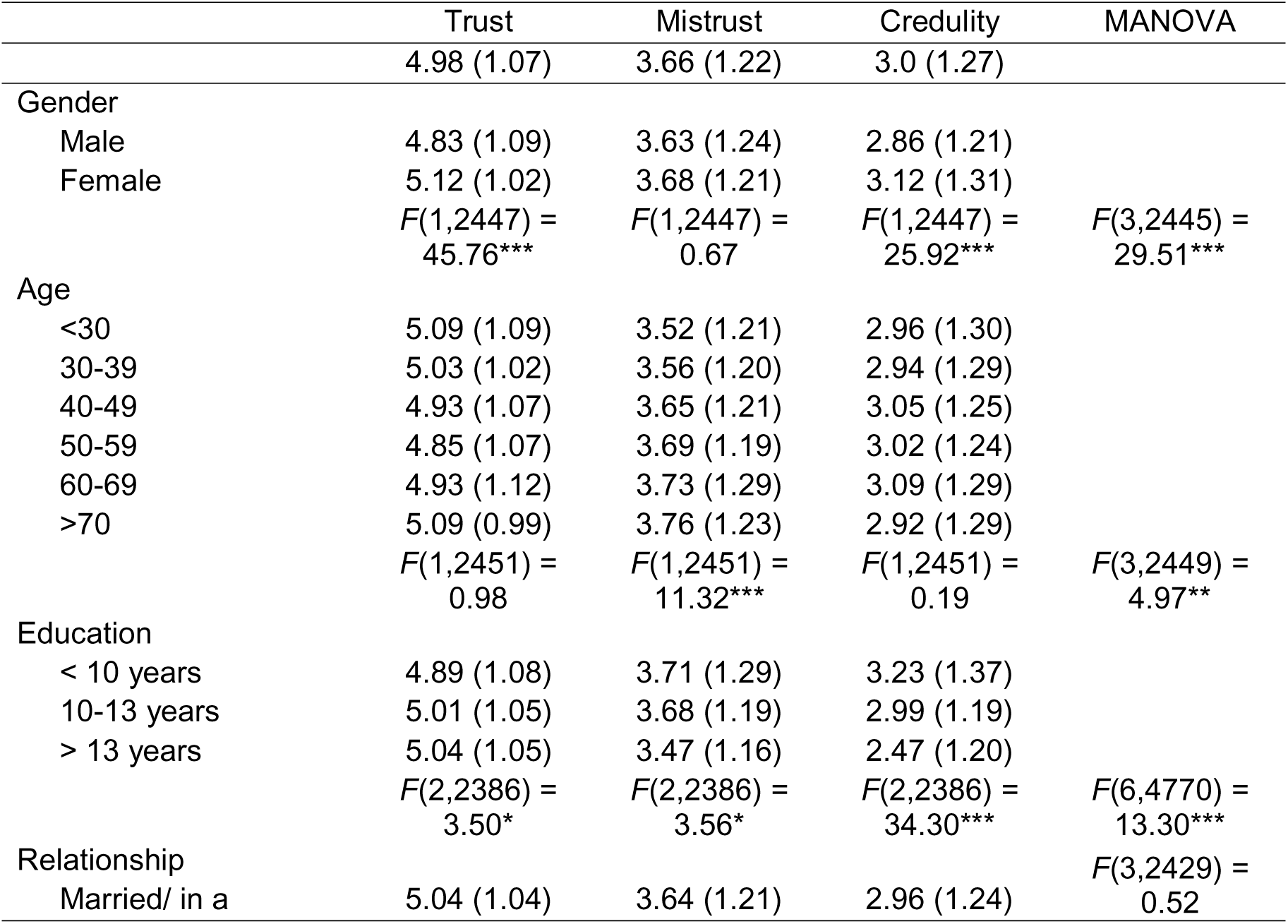
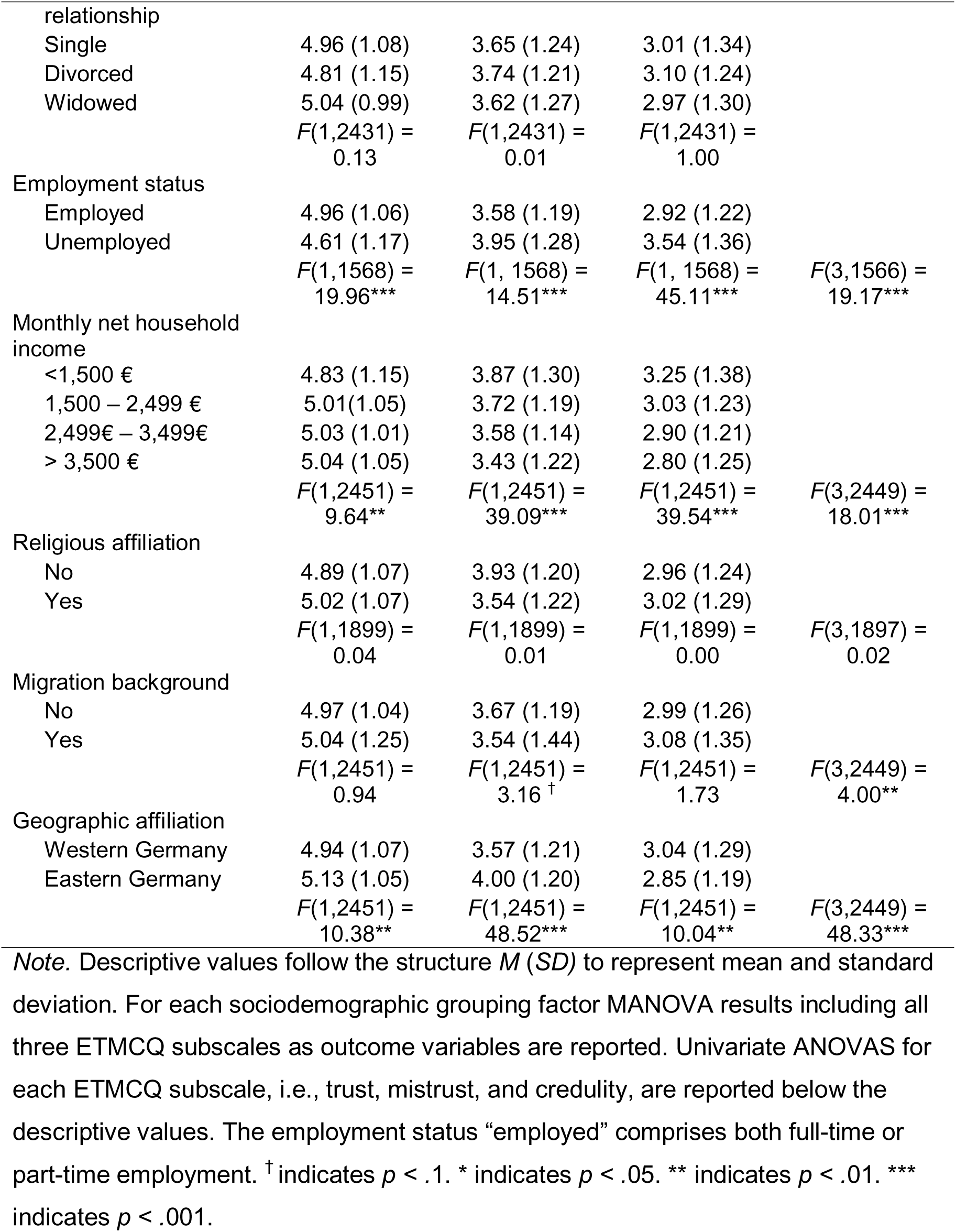
Sociodemographic group differences in epistemic stance.

### Discriminant and Convergent Validity of the ETMCQ

Table 3 presents Spearman correlations between the ETMCQ subscales and constructs related to adversity, symptomatology, functioning, and attachment styles.

A False Discovery Rate (FDR) correction was applied to account for multiple comparisons.

**Table 3.** Correlations of ETMCQ subscales with psychological questionnaire scores (adversity: ACE, symptomatology: ITQ, PHQ-9, GAD-7, PSS-4 and personality functioning: OPD-SQS, ECR-R, AAR)

| Questionnaire | Trust | Mistrust | Credulity |
| --- | --- | --- | --- |
| ETMCQ Mistrust | -.16*** |  |  |
| ETMCQ Credulity | -.14*** | .57*** |  |
| ACE | -0.11*** | 0.18*** | 0.25*** |
| ITQ |  |  |  |
| PTSD | -0.03 | 0.16*** | 0.24*** |
| DSO | -0.18*** | 0.31*** | 0.38*** |
| OPD-SQS |  |  |  |
| self-perception | -0.15*** | 0.37*** | 0.46*** |
| interpersonal contact | -0.18*** | 0.41*** | 0.48*** |
| relationship model | -0.09*** | 0.52*** | 0.48*** |
| total | -0.16*** | 0.51*** | 0.54*** |
| PHQ-9 | -0.11*** | 0.27*** | 0.35*** |
| GAD-7 | -0.08*** | 0.27*** | 0.32*** |
| PSS-4 | -0.18*** | 0.22*** | 0.38*** |
| ECR-R |  |  |  |
| avoidance | -0.19*** | 0.17*** | 0.18*** |
| anxiety | -0.07*** | 0.19*** | 0.25*** |
| AAR |  |  |  |
| secure | 0.26*** | -0.18*** | -0.26*** |
| excessive dependency | -0.19*** | 0.25*** | 0.44*** |
| interpersonal ambivalence | -0.24*** | 0.32*** | 0.38*** |
| compulsive care-giving | -0.04* | 0.11*** | 0.22*** |
| rigid self-control | 0.05* | 0.12*** | -0.05** |
| defensive separation | -0.23*** | 0.25*** | 0.21*** |
| emotional detachment | -0.35*** | 0.28*** | 0.24*** |

*Note.* ETMCQ = Epistemic Trust, Mistrust and Credulity Questionnaire, ITQ PTSD = International Trauma Questionnaire post-traumatic stress disorder subscale, ITQ DSO = International Trauma Questionnaire disturbance in self organization subscale, ACE = Adverse Childhood Experiences, OPD-SQS = Short Operationalised Psychodynamic Diagnostics Questionnaire – Structural Axis, PHQ = Patient Health Questionnaire, GAD = Generalised Anxiety Disorder Questionnaire, PSS = Perceived Stress Scale, ECR-R = Revised Experience in Close Relationships Questionnaire, AAR = Adult Attachment Ratings. All *p*-values were adjusted using a False Discovery Rate (FDR) correction to account for multiple comparisons. \* indicates $p < .05$ . \*\* indicates $p < .01$ . \*\*\* indicates $p < .001$ .

Significant negative correlations were observed between Trust and maltreatment, difficulties in self-organisation (as part of complex PTSD), personality functioning, levels of anxiety and depression, perceived stress, and avoidant and anxious attachment styles. In contrast, both Epistemic Mistrust and Credulity showed positive correlations with these variables, indicating that higher endorsement of Mistrust and Credulity was associated with greater symptom severity, reduced personality functioning, and higher levels of insecure attachment. Notably, Credulity yielded slightly stronger correlation coefficients than Mistrust.

To assess the discriminant validity of the ETMCQ with regard to depression and anxiety, we examined differences in epistemic stance between individuals above and below the clinical cut-offs for PHQ-9 (depression: ≥ 10) and GAD-7 (anxiety: ≥ 10). Table 4 summarises the results of Welch’s *t*-tests. Mistrust and Credulity emerged as significant discriminators between anxiety and depression caseness, with individuals above the cut-offs exhibiting increased levels of both. Effect sizes were slightly higher for Credulity than for Mistrust. The Trust subscale only differentiated those above and below the depression cut-off, showing a small effect size, while it did not significantly discriminate anxiety.

**Table 4.** Differences in epistemic stance between PHQ-9 and GAD-7 case groups.

| Comparison | Subscale | No Case<br>*M* (*SD*) | Case<br>*M* (*SD*) | *t*-test | *p*-<br>value | Cohen's<br>*d* |
| --- | --- | --- | --- | --- | --- | --- |
| PHQ-9<br>no case (*n* =<br>2360);<br>case (*n* = 153) | Trust | 5.01 (1.06) | 4.62 (1.14) | *t*(169.63) =<br>4.14 | < .001 | 0.72 |
|  | Mistrust | 3.60 (1.20) | 4.49 (1.27) | *t*(170.40) = -<br>8.42 | < .001 | 0.72 |
|  | Credulity | 2.93 (1.24); | 4.06 (1.28) | *t*(169.97) = -<br>10.57 | < .001 | 0.90 |
| GAD-7<br>no case (*n* =<br>2403);<br>case (*n* = 108) | Trust | 4.99 (1.06); | 4.73 (1.24) | *t*(114.10) =<br>2.19 | .031 | 0.23 |
|  | Mistrust | 3.61 (1.20); | 4.61 (1.34) | *t*(113.86) = -<br>7.63 | < .001 | 0.79 |
|  | Credulity | 2.95 (1.24); | 4.09 (1.43) | *t*(114.42) = -<br>8.13 | < .001 | 0.85 |

### Childhood Adversity, Epistemic Trust, and Mental Health Symptoms

To examine whether the effects of childhood adversity (ACE) on mental health symptoms were mediated by epistemic stance, we tested four mediation models. For each model, estimates, average causal mediation effects (ACME), and 95% confidence intervals (CI) were computed using 1,000 bootstrapped simulations. The mental health outcomes assessed included depressive symptoms (PHQ-9), anxiety symptoms (GAD-7) and perceived stress (PSS-4).

The analyses consistently showed that childhood adversity was associated with higher levels of symptomatology. This effect was partially mediated by epistemic stance across all four outcomes. The pattern revealed that childhood adversity was linked to lower Trust and higher Mistrust and Credulity, which in turn affected mental health outcomes.

For depressive symptoms, both Mistrust (ACME = 0.01, 95% CI [0.01, 0.02]) and Credulity (ACME = 0.05, 95% CI [0.04, 0.07]) acted as significant mediators, contributing to higher symptom severity. Trust, however, did not emerge as a significant protective factor (ACME = 0.00, 95% CI [0.00, 0.01]).

Similarly, a significant proportion of the effect of childhood adversity on anxiety symptoms was explained by increased Mistrust (ACME = 0.02, 95% CI [0.01, 0.03]) and Credulity (ACME = 0.05, 95% CI [0.03, 0.06]), whereas Trust again did not act as a mediator (ACME = 0.00, 95% CI [0.00, 0.01]).

Lastly, the effect of childhood adversity on perceived stress was mediated by both higher levels of Credulity (ACME = 0.08, 95% CI [0.06, 0.10]) and, in contrast to other outcomes, higher Trust (ACME = 0.01, 95% CI [0.01, 0.02]). No significant association with Mistrust was detected for perceived stress (ACME = 0.00, 95% CI [- 0.01, 0.01]).

The proportion of the total effect of childhood adversity that was mediated by epistemic stance ranged from 17.5% for depressive symptoms to 32.1% for perceived stress. For an illustration, see Figures 2–5 (estimations of the indirect effects are provided in the supplementary information).

**Figure 2.**
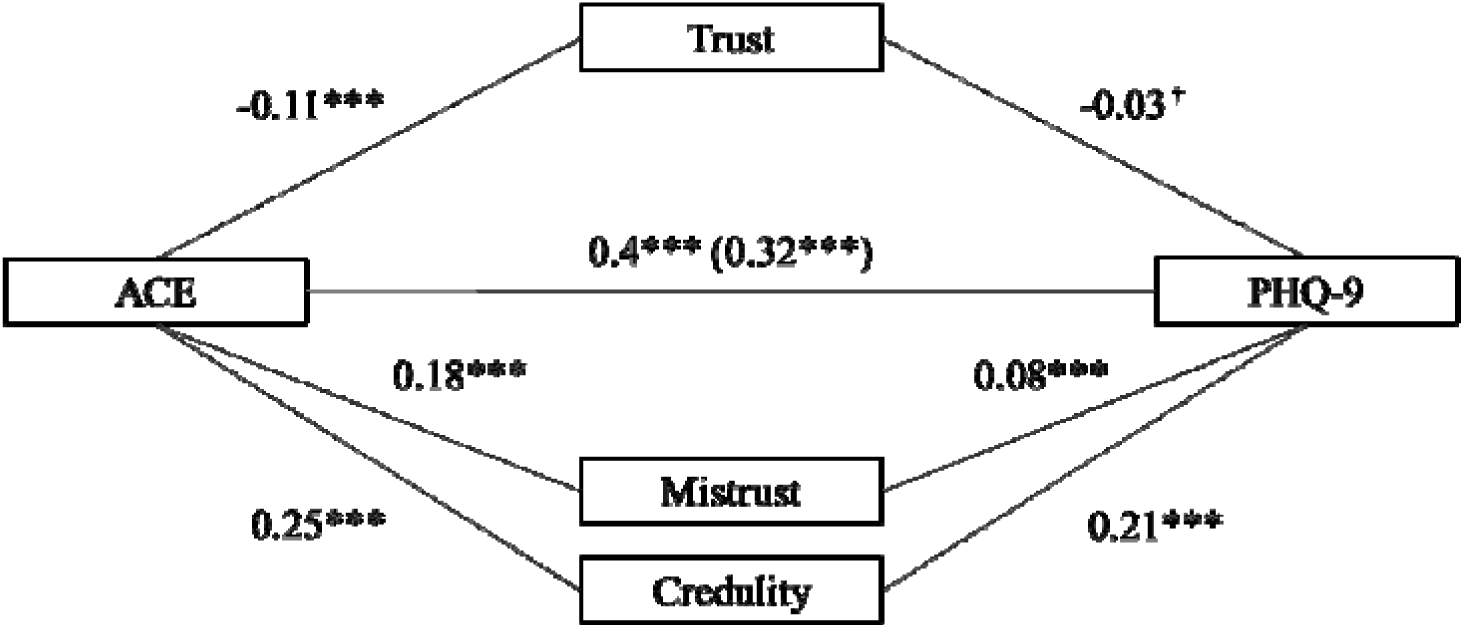
Mediation of ACE on PHQ-9 across ETMCQ. *Note.* Standardized estimates were computed using 1000 bootstrapped simulations. The estimate for the direct effect of ACE on PHQ-9 controlling for the ETMCQ mediators is in parentheses. *R^2^* = .23; *F*(4,2514) = 186.68; *p* = < .001. * indicates *p < .*05. *** indicates *p < .*001.

**Figure 3.**
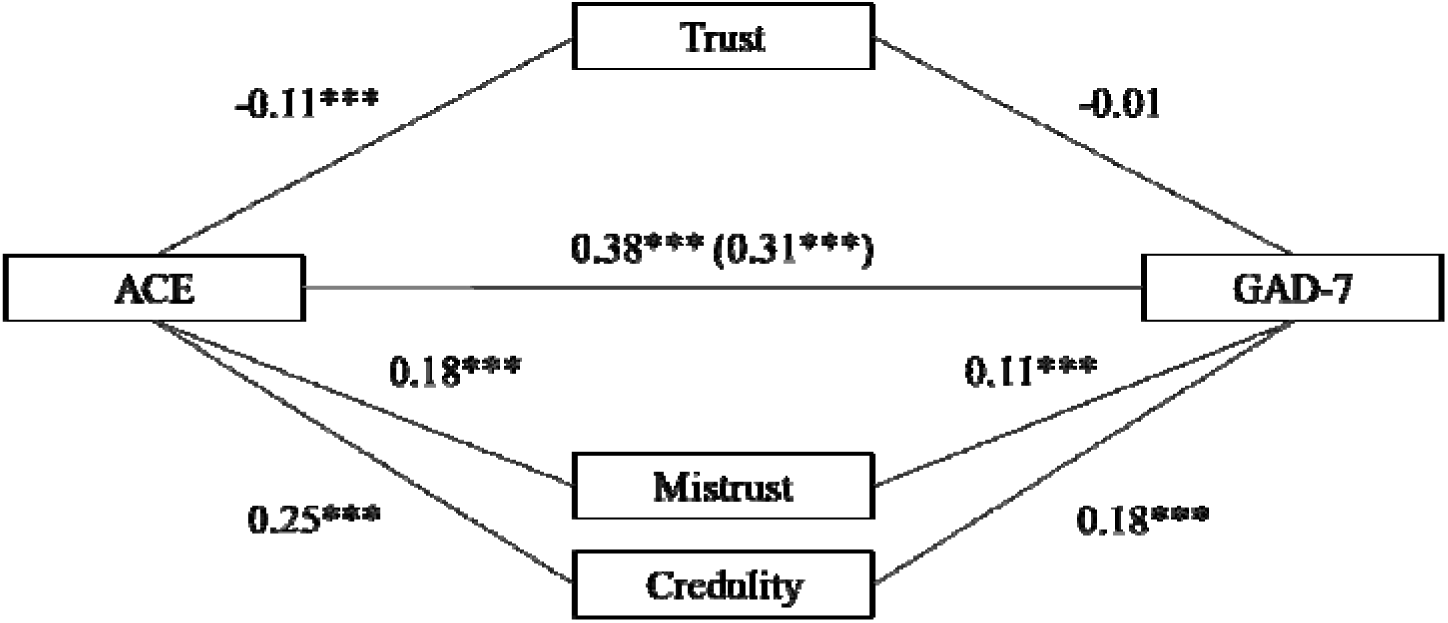
Mediation of ACE on GAD-7 across ETMCQ. *Note.* Standardized estimates were computed using 1000 bootstrapped simulations. The estimate for the direct effect of ACE on GAD-7 controlling for the ETMCQ mediators is in parentheses. *R^2^* = .21; *F*(4,2514) = 162.83; *p* = < .001. *** indicates *p < .*001.

**Figure 4.**
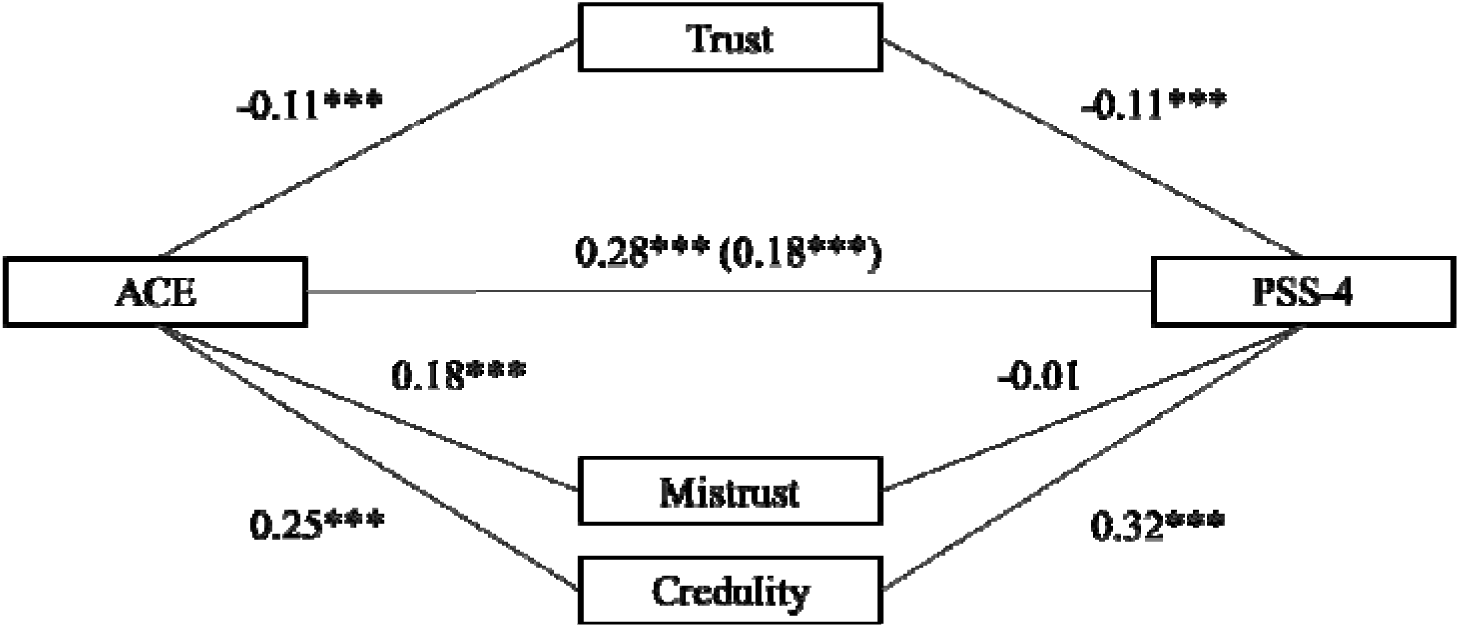
Mediation of ACE on PSS-4 across ETMCQ. *Note.* Standardized estimates were computed using 1000 bootstrapped simulations. The estimate for the direct effect of ACE on PSS-4 controlling for the ETMCQ mediators is in parentheses. *R^2^* = .19; *F*(4,2514) = 147.42; *p* = < .001. *** indicates *p < .*001.

### Differences in Epistemic Stance between Attachment Groups

Participants were grouped according to their attachment patterns using a median split on the avoidance and anxiety scales of the ECR-R. This resulted in four attachment categories: low avoidance/low anxiety (*n* = 1,066), low avoidance/high anxiety (*n* = 574), high avoidance/low anxiety (*n* = 143), and high avoidance/high anxiety (*n* = 497). A MANOVA was conducted to assess differences in epistemic stance between these groups, with all three ETMCQ subscales as outcome variables. The results are presented in Table 5. Figures 6–8 show bar plots representing pairwise comparisons between attachment groups, conducted using pairwise *t*-tests with a False Discovery Rate (FDR) correction for multiple comparisons.

**Figure 6.**
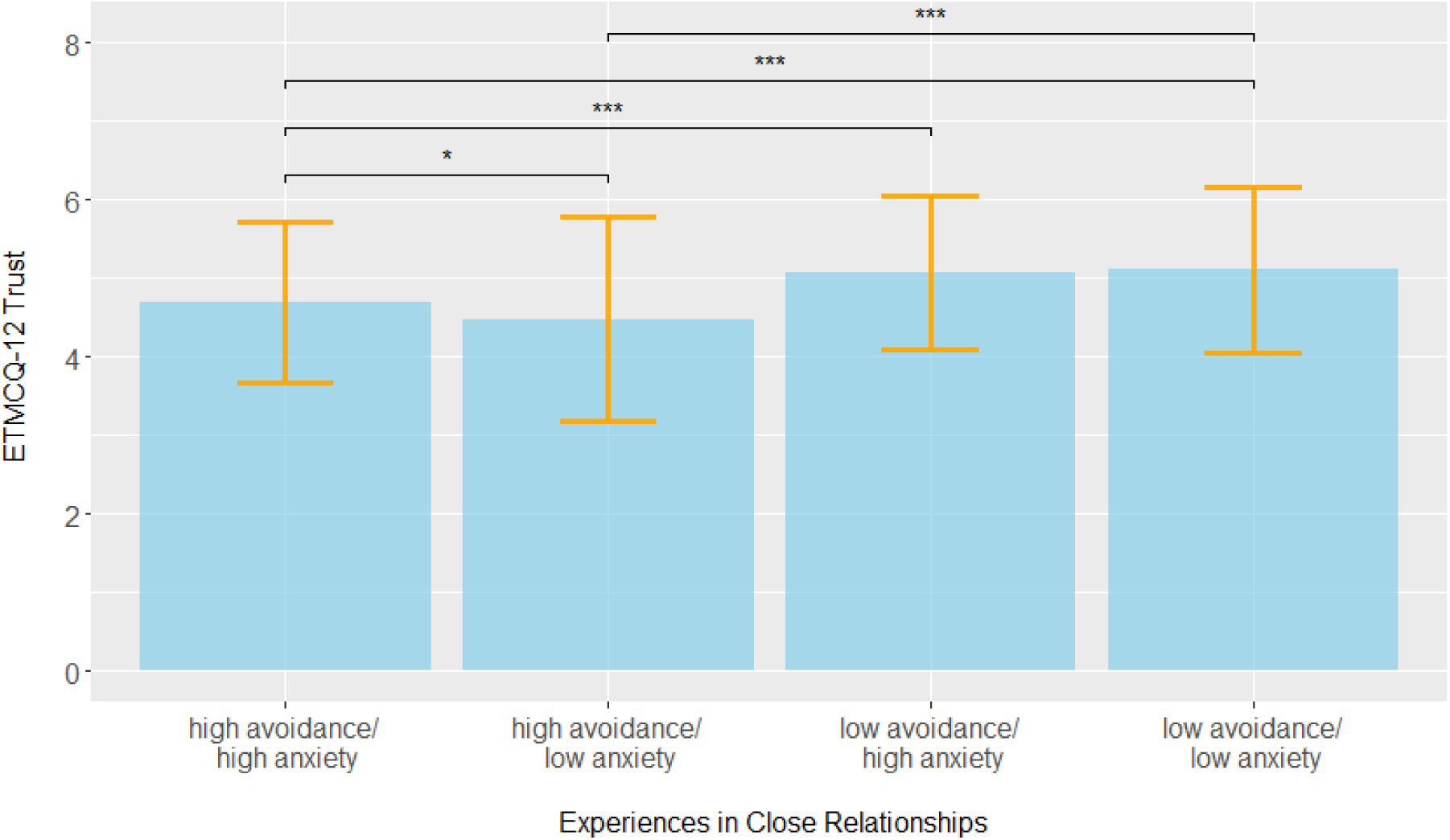
ECR-R group differences in epistemic trust. *Note.* All *p*-values were adjusted using a False Discovery Rate (FDR) correction to account for multiple comparisons. * indicates *p < .*05. ** indicates *p < .*01. *** indicates *p < .*001.

**Figure 7.**
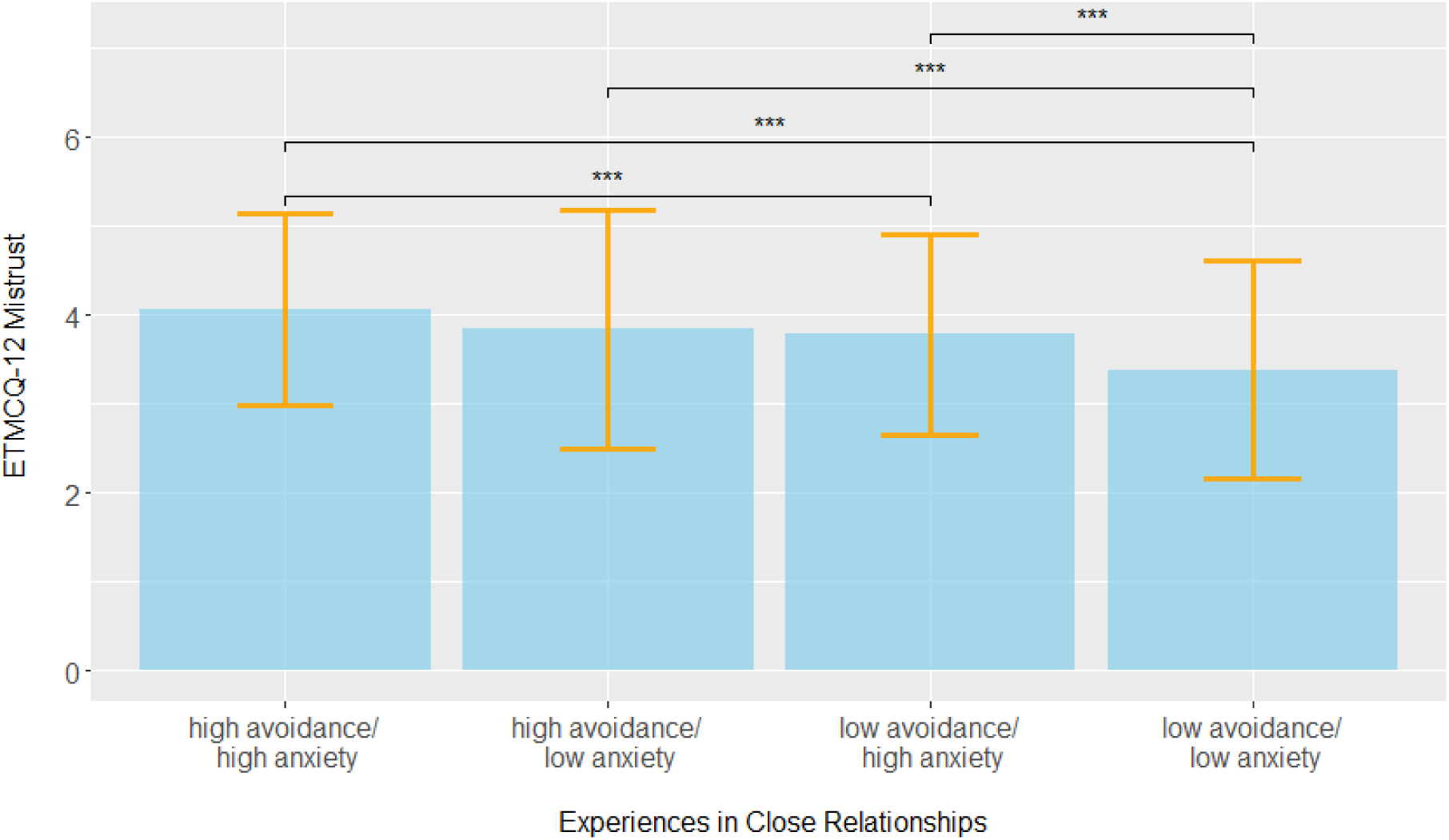
ECR-R group differences in epistemic mistrust. *Note.* All *p*-values were adjusted using a False Discovery Rate (FDR) correction to account for multiple comparisons. * indicates *p < .*05. ** indicates *p < .*01. *** indicates *p < .*001.

**Figure 8.**
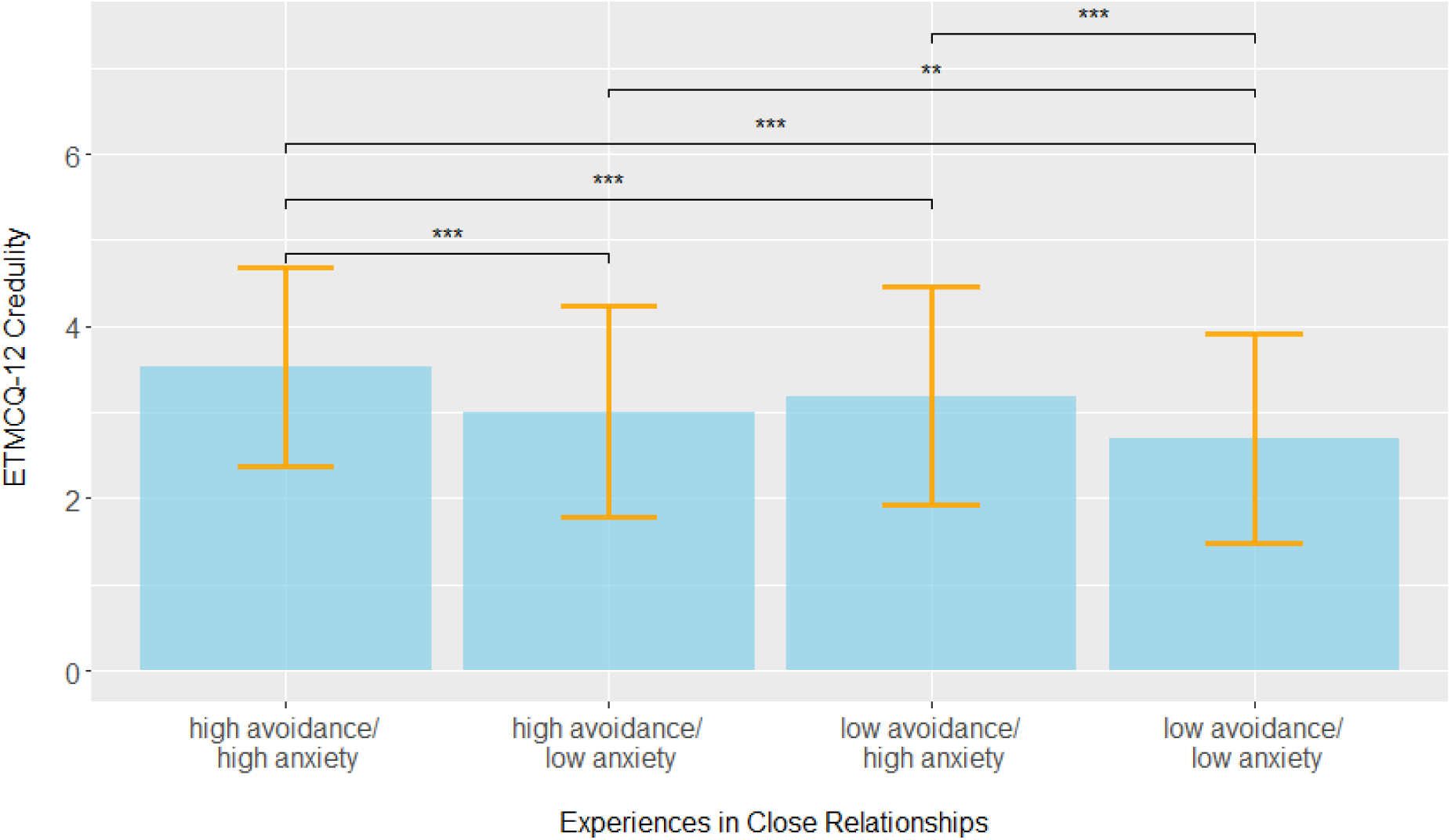
ECR-R group differences in epistemic credulity. *Note.* All *p*-values were adjusted using a False Discovery Rate (FDR) correction to account for multiple comparisons. * indicates *p < .*05. ** indicates *p < .*01. *** indicates *p < .*001.

**Table 5.** ECR-R group differences in epistemic stance.

|  | Trust | Mistrust | Credulity | MANOVA |
| --- | --- | --- | --- | --- |
| low avoidance/<br>low anxiety | 5.11 (1.06) | 3.38 (1.23) | 2.70 (1.22) |  |
| low avoidance/<br>high anxiety | 5.07 (0.98) | 3.78 (1.13) | 3.19 (1.27) |  |
| high avoidance/<br>low anxiety | 4.48 (1.30) | 3.84 (1.35) | 3.01 (1.23) |  |
| high avoidance/<br>high anxiety | 4.69 (1.03) | 4.06 (1.07) | 3.53 (1.15) |  |
| | $F(3,2223) = 29.58.76^{***}$ | $F(3,2223) = 41.12^{***}$ | $F(3,2223) = 57.57^{***}$ | $F(9,6669) = 28.13^{***}$ |

*Note.* Descriptive values follow the structure $M(SD)$ to represent mean and standard deviation. MANOVA results including all three ETMCQ subscales as outcome variables are reported. Univariate ANOVAs for each ETMCQ subscale, i.e., trust, mistrust, and credulity, are reported below the descriptive values. \*\*\* indicates $p < .001$ .

The findings indicate that individuals characterised by low attachment avoidance and anxiety (approximating attachment security) exhibited the highest levels of Trust and the lowest levels of Mistrust and Credulity. In contrast, those with high levels of both avoidance and anxiety (approximating fearful attachment) showed the opposite pattern, with lower Trust and higher endorsement of Mistrust and Credulity.

Furthermore, there is some convergence of these patterns of associations when comparing ECR-R results with the Adult Attachment Ratings (AAR) approach (bottom of table 3). Most notably, secure attachment measured this way is positively correlated with secure attachment with the oppositve association emerging for both types of epistemic disruption. The reverse pattern was found for correlations with interpersonal ambivalence, compulsive caregiving, defensive separation and emotional detachment. Higher expression of rigid self control was associated with higher Trust and Credulity and lower Mistrust.

### Establishing Cut-offs for the ETMCQ in Relation to Anxious and Depressive Symptomatology

For the Trust subscale, individuals scoring below the cut-off were more likely to exhibit depressive (PHQ-9) or anxious (GAD-7) symptoms, while for the Mistrust and Credulity subscales, individuals scoring above the cut-off were more likely to show depression or anxiety. However, due to the relatively small number of cases within this predominantly healthy sample, the Area Under the Curve (AUC) scores for Trust in relation to both depression and anxiety remained below the conventional threshold of 0.70. This suggests that epistemic disruption, rather than low levels of Trust, might be a more sensitive indicator of psychopathology in this context.

## Discussion

### Factor Structure

The current study validated the German version of the ETMCQ by examining its factor structure and extending previous findings related to its convergent and discriminant validity in a representative sample of over 2,000 participants. Our findings support the established three-factor structure of the instrument, which distinguishes between trusting, mistrusting, and credulous predispositions in the acquisition of social information (Campbell et al., 2021; Liotti et al., 2023; Asgarizadeh et al., 2023; Greiner et al., 2024). To achieve the best model fit, the German version required reducing the original 15-item scale to a 12-item version, consistent with preliminary results from a non-representative German sample (Weiland et al., 2024).

The EFA indicated that some items intended to capture mistrust or credulity loaded onto a common factor of epistemic disruption, which prompted the testing of a two-factor solution in the CFA. This result warrants replication in further non-clinical samples. Pearson correlations between the three ETMCQ factors in our sample differed from previous studies by Campbell and Liotti, particularly with higher correlations between credulity and mistrust (r = 0.57). This suggests that these two seemingly opposing predispositions may largely share a common underlying cause— epistemic disruption. The latter may suggest that rational judgements of trustworthy versus untrustworthy soulces cannot be made which may lead people to be more predispoosed to be hypervigilant at the same time as being somewhat at a loss when judgements of trustworthyness need to be made. However, caution is advised, as only three items contributed to the credulity subscale, and item reduction (or linguistic nuances in the German version) may have exaggerated the relationship between the two factors. Nevertheless, this finding aligns with the original ETMCQ validation, where mistrust and credulity were significantly correlated (r = .47, p < .001).

Additionally, we found significant negative associations between trust and both mistrust and credulity, contributing to the complex picture of how these factors relate across different questionnaire versions.

Future research should focus on refining the instrument and incorporating recent advancements in the ETMCQ’s psychometric properties, as demonstrated by Campbell et al.’s (2024, under review) revision of the questionnaire.

### Relationships with Demographic Variables

Using a representative sample allowed for an in-depth analysis of the relationships between epistemic stance and demographic characteristics. Consistent with Campbell et al. (2021), but differing from Liotti et al. (2023), we found that men and women showed significant differences in both the Trust and Credulity subscales, with women scoring higher than men on both. Due to design limitations, participants identifying as non-binary or outside of male/female categories were not included in the analyses, which is unfortunate and highlights the need for future research to incorporate these individuals’ epistemic stance.

Additional associations with demographic factors emerged, indicating that higher educational attainment was linked to significantly lower epistemic disruption.

Conversely, unemployment and lower household income were associated with reduced trust and higher epistemic disruption. Living in the eastern part of Germany (former German Democratic Republic) was linked with higher levels of trust, mistrust, and lower credulity (with other demographic variables controlled for). Individuals with a migration background exhibited higher trust and lower epistemic disruption, though these effects were relatively small.

These findings suggest that cultural, societal, and economic factors—largely underexplored in the context of social communication—may play a role in shaping epistemic stance. These factors, potentially tied to experiences of marginalisation, epistemic isolation, and reduced access to resources, are often exacerbated by intersectionality (Campbell & Allison, 2023; Mau, Lux & Heide, 2024). Marginalisation may predict shame which leads to the withdrawal from the community of those sharing knowledge. The feeling that marginalised people have of exclusion and loneliness/isolation makes them find common cause with others who feel the same way. This then may contribute to the lowering of epistemic vigilance in relation to these individuals (including the potential for exploitation by them) which accounts for part of the paradoxical association between mistrust and credulity. To better understand the interplay of these variables, future research should investigate these influences directly, particularly in high-risk or disadvantaged samples.

### Convergent and Discriminant Validity

In line with our second set of hypotheses, based on Campbell et al.’s findings, the correlations between the ETMCQ subscales and adverse childhood experiences, psychopathology, attachment, and personality functioning were largely confirmed. Building on previous literature (Benzi et al., 2023; Greiner et al., 2024; Kampling et al., 2022; Liotti et al., 2023; Tironi et al., 2024), early adversity in this study was linked with epistemic disruption and reduced epistemic trust. Notably, the strength of the negative correlation between Trust and psychopathology was weaker (or absent in the case of PTSD symptoms) compared to the relationship between epistemic disruption (Mistrust and Credulity) and symptom severity. Interestingly, higher correlation coefficients in the same directions where found the disturbance in self- organistion subscale of the ITQ than for PTSD difficulites which lends support to the notion that the former might be more affected by epistemic disruption due to the lack of trauma processing from a mentalizing social environement (Smits et al., 2024).

When predicting anxiety and depression cases (Table 6), the ETMCQ subscale perform reasonably well apart from Trust with Credulity being superior to Mistrust in terms of area under the curve predictions.

**Table 6.** Predicting depression and anxiety cases with epistemic stance.

|  | optimal cut-off | specificity | sensitivity | AUC |
| --- | --- | --- | --- | --- |
| Depression |  |  |  |  |
| Trust | 4.70 | 0.64 | 0.52 | 0.60 |
| Mistrust | 4.50 | 0.78 | 0.50 | 0.70 |
| Credulity | 3.13 | 0.56 | 0.78 | 0.73 |
| Anxiety |  |  |  |  |
| Trust | 3.90 | 0.86 | 0.28 | 0.57 |
| Mistrust | 4.50 | 0.78 | 0.56 | 0.71 |
| Credulity | 3.63 | 0.68 | 0.66 | 0.73 |

Trust did not mediate the relationship between early adversity and anxiety and depression, with the exception of perceived stress, where a buffering effect of trust was found. This may reflect limitations in the construct validity of the Trust subscale. However, Campbell et al. (2021) suggested that epistemic trust may represent a “default mode” of social functioning, i.e. the notion that increasing trust beyond a certain point is not adding to protection from pathology because it is feeding into credulity and epistemic disruption and thus providing no additional clinical benefit beyond the average level.

As expected, the correlation coefficients were stronger for Mistrust and Credulity, reinforcing the idea that higher levels of early adversity lead to an "epistemic dilemma" (Campbell & Fonagy, 2022). This dilemma likely drives individuals to oscillate between epistemic credulity and mistrust, impairing their ability to engage in appropriate knowledge transfer. As Campbell et al. (2021) posited, such early adversities might generate “epistemic petrification” or a loss of appropriate vigilance, with individuals alternating between mistrust and credulity. Over time, this leads to an exhaustion of the epistemic system, reducing the individual’s ability to benefit from social learning and access the salutogenic factors that might otherwise be available from the environment (Fonagy et al., 2017; Nolte et al., 2023). Clinically, this restricted social learning, often exacerbated by experiences of abuse or neglect, further diminishes the individual’s ability to assess trustworthiness and maintain a stable epistemic stance into adulthood (Luyten, Campbell & Fonagy, 2020).

It is important to note that these associations and mediation patterns were observed in a community sample. While adverse childhood experiences (ACEs) were present, their prevalence and severity were lower than in high-risk populations. Therefore, further validation is needed from clinical samples with higher levels of early adversity, where the impact of dose, onset, and type of maltreatment can be more thoroughly explored. The cross-sectional nature of this study also precluded examination of factors like mentalizing, which might mitigate the disruption of social learning and the establishment of appropriate epistemic trust (Fonagy et al., 2014; Campbell et al., 2021; Nolte et al., 2023).

It is noteworthy, that in this sample, there were high correlation coefficients between the ETMCQ subscales and the short OPD-SQ total scores indicating largely overlapping constructs. This deserves further attention in psychometrics assessment in future research.

Our post-hoc exploration of the relationship between attachment styles (secure, fearful, dismissive, and preoccupied, based on median splits of ECR-R scores derived on the core item for each ECR-R subscale) and epistemic stance indicated that different attachment patterns are linked with distinct epistemic profiles.

Consistent with Campbell et al.’s (2021) findings, insecure attachment styles were positively associated with both Credulity and Mistrust – with the combination of high avoidance and high anxiety being associated with highlest levels of mistrust and credulity, while secure attachment was linked to the lowest levels of epistemic disruption and the highest levels of Trust. These results support the developmental framework suggesting that secure attachment facilitates the formation of basic trust in infancy, fostering an epistemic stance conducive to interpersonal learning (Luyten et al., 2020). Secure attachment figures, along with mentalizing relationships or social systems, may contribute to an individual’s capacity to assume basic trust in others (Zeegers et al., 2017). The findings further support a developmental notion according to which people with high avoidance scores adopt a secondary attachment strategy of deactivating attachment concerns while those with anxious aviadance are hyperactivating attachment with the latter predicting credulity while deactivation would be linked with mistrust. These seems to be the case in our data although high levels of attachment anxiety were also found in those expressing high levels of mistrust. As expected, a largely similar set of associations was found for correlations between the three aspects of epistemic stance and the subsclales of the Adult Attachment Ratings systems, a pragmatic way to assess attachment characteristics in larger surveys.

To deepen our understanding of the interplay between adversity, attachment strategies, epistemic stance, social functioning, and psychopathology, future research should explore risk and resilience factors, particularly the role of being mentalized and recognised as social agents. Longitudinal studies are needed to disentangle the bi-directional relationships between these variables.

### Limitations

Several limitations should be considered when interpreting the findings of this study. First, all variables were assessed using self-report measures, introducing the possibility of shared methods variance. Additionally, childhood adversity was assessed retrospectively, which may have impacted the accuracy of recall. The study design did not permit the assessment of test-retest reliability for the ETMCQ, limiting our ability to evaluate the temporal stability of the instrument.

While the sample was drawn from a representative German population, it likely included individuals with mental health difficulties; however, no clinical corroboration of diagnostic status was obtained, restricting our ability to draw conclusions about epistemic stance in clinical populations. Moreover, the cross-sectional nature of the study prevents any causal inferences; the findings represent associations between variables rather than causal relationships.

Another limitation relates to the lack of experimental evidence to further support the validity of the instrument. Social learning processes in real-world settings are likely more complex, with additional mechanisms involved that cannot be fully captured by self-reported predispositions. Lastly, our data originate from a WEIRD (Western, Educated, Industrialized, Rich, Democratic) population (Henrich, 2022). While the use of a representative German sample is a strength, the results may not be generalisable to other populations or cultural contexts.

### Conclusion

Epistemic trust is essential for the assimilation of new information, enabling individuals to engage adaptively with their changing social environment (Fonagy & Allison, 2014). A key element of this process is the ability to adopt an appropriate epistemic stance, assessing when to trust or mistrust others. This involves accurately interpreting a communicator’s intentions, assessing the accuracy and relevance of the information, and weighing these factors against one’s existing knowledge.

Drawing on a representative sample, our findings provide support for the psychometric robustness of the German version of the ETMCQ. However, further research is needed to refine and improve the instrument, ensuring it captures the complexities of epistemic stance across different contexts and populations.

## Data Availability

All data produced in the present study are available upon reasonable request to the authors

## Supplements

**Table 1.**
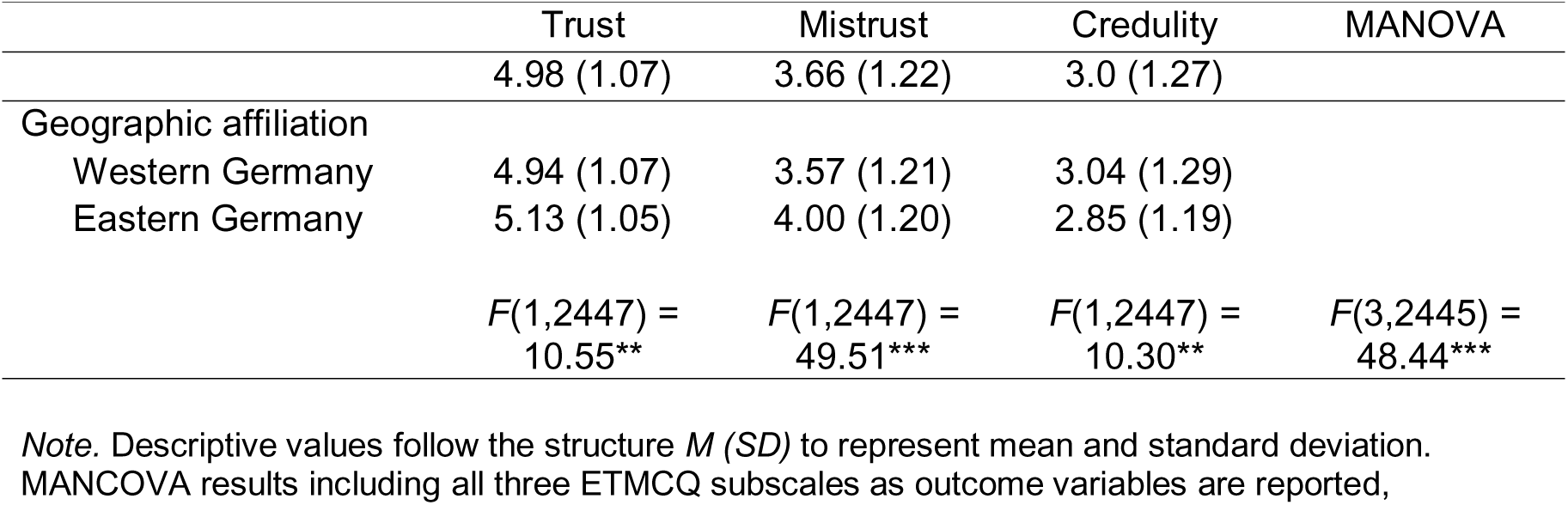
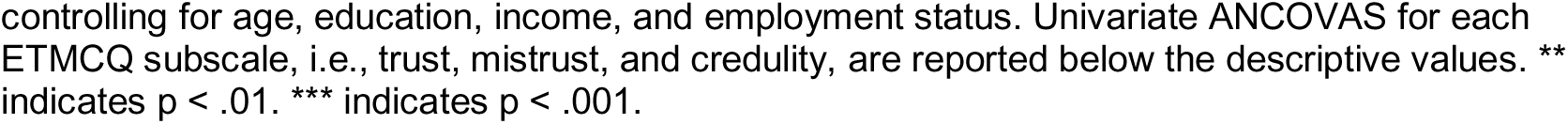
Effects of geographic affiliation on ETMCQ controlled for counfounding demographic variables.

|  | Trust | Mistrust | Credulity | MANOVA |
| --- | --- | --- | --- | --- |
|  | 4.98 (1.07) | 3.66 (1.22) | 3.0 (1.27) |  |
| Geographic affiliation |  |  |  |  |
| Western Germany | 4.94 (1.07) | 3.57 (1.21) | 3.04 (1.29) |  |
| Eastern Germany | 5.13 (1.05) | 4.00 (1.20) | 2.85 (1.19) |  |
| | $F(1,2447) = 10.55^{**}$ | $F(1,2447) = 49.51^{***}$ | $F(1,2447) = 10.30^{**}$ | $F(3,2445) = 48.44^{***}$ |

*Note.* Descriptive values follow the structure $M (SD)$ to represent mean and standard deviation. MANCOVA results including all three ETMCQ subscales as outcome variables are reported,
controlling for age, education, income, and employment status. Univariate ANCOVAs for each ETMCQ subscale, i.e., trust, mistrust, and credulity, are reported below the descriptive values. \*\* indicates $p < .01$ . \*\*\* indicates $p < .001$ .

**Table 2.** Indirect effect estimates from mediation models of ACE on mental health outcomes across ETMCQ.

|  | Indirect effect | CI lower | CI upper |
| --- | --- | --- | --- |
| PHQ-9 |  |  |  |
| Trust | 0.00 | 0.00 | 0.01 |
| Mistrust | 0.01 | 0.01 | 0.02 |
| Credulity | 0.05 | 0.04 | 0.07 |
| Total | 0.07 | 0.06 | 0.09 |
| GAD-7 |  |  |  |
| Trust | 0.00 | 0.00 | 0.01 |
| Mistrust | 0.02 | 0.01 | 0.03 |
| Credulity | 0.05 | 0.03 | 0.06 |
| Total | 0.07 | 0.05 | 0.08 |
| PSS-4 |  |  |  |
| Trust | 0.01 | 0.01 | 0.02 |
| Mistrust | 0.00 | -0.01 | 0.01 |
| Credulity | 0.08 | 0.06 | 0.10 |
| Total | 0.09 | 0.08 | 0.11 |

*Note.* Standardized indirect effect estimates and confidence intervals (CIs) were computed using 1000 bootstrapped simulations. Effects are shown for each mediator and in total for the four mediation outcomes, respectively.

## References

Backhaus, K., Erichson, B., Plinke, W., & Weiber, R. (2016). Mutlivariate Analysemethoden. Eine anwendungsorientierte Einführung. Berlin, Heidelberg: Springer Gabler.

Beck U, Giddens A, Lash S. Reflexive modernization: Politics, tradition and aesthetics in the modern social order. Stanford University Press; 1994.

Benzi, Ilaria Maria Antonietta, et al. "Different epistemic stances for different traumatic experiences: implications for mentalization." Research in Psychotherapy: Psychopathology, Process, and Outcome 26.3 (2023).

Bowlby J. The bowlby-ainsworth attachment theory. Behavioral and Brain Sciences. 1979;2(4):637–8.

Campbell, C., & Fonagy, P. (2022). Bifocalism is in the eye of the beholder: Social learning as a developmental response to the accuracy of others’ mentalizing. Behavioral and Brain Sciences, 45, e254, Article e254. 10.1017/S0140525X22001297

Campbell, C., Delamain, H., Saunders, R., Tanzer, M., Milesi, A., Nolte, T., Allison, E., Luyten, P., & Fonagy, P. (Under submission). Development and Validation of the Epistemic Trust, Mistrust and Credulity Questionnaire – Revised (ETMCQ-R).

Cloitre, M.; Shevlin, M.; Brewin, C.R.; Bisson, J.I.; Roberts, N.P.; Maercker, A.; Karatzias, T.; Hyland, P. The International Trauma Questionnaire: Development of a self-report measure of ICD-11 PTSD and complex PTSD. Acta Psychiatr. Scand. 2018, 138, 536–546.

Csibra, G., & Gergely, G. (2006a). Social learning and social cognition: The case for pedagogy. In M. H. Johnson & Y. Munakata (Eds.), Processes of change in brain and cognitive development. Attention and Performance XXI (pp. 249–274). Oxford University Press.

Gergely, G., & Király, I. (2019). Natural pedagogy of Social Emotions.. In F. Clements & D. Dukes (Eds.), Affective Social Learning: Conceptualizing the Social Transmission of Value (pp. 87-114). Cambridge University Press. 10.1017/9781108661362

Gergely, G., & Watson, J. S. (1996). The social biofeedback theory of parental affect- mirroring: The development of emotional self-awareness and self-control in infancy. International Journal of Psychoanalysis, 77(Pt 6), 1181–1212. http://www.ncbi.nlm.nih.gov/pubmed/9119582

Gergely, G., & Csibra, G. (2005). The social construction of the cultural mind: Imitative learning as a mechanism of human pedagogy. Interaction Studies, 6, 463–481. 10.1075/is.6.3.10ger

Gergely, G., & Csibra, G. (2006b). Sylvia’s recipe: Human culture, imitation, and pedagogy. In N. J. Enfield & S. C. Levinson (Eds.), Roots of human sociality: Culture, cognition, and human interaction (pp. 229–255). Berg Press.

O’Connor, B.P. SPSS and SAS programs for determining the number of components using parallel analysis and velicer’s MAP test. Behav. Res. Methods Instrum. Comput. 2000, 32, 396–402.

Ehrenthal, J.C.; Dinger, U.; Schauenburg, H.; Horsch, L.; Dahlbender, R.W.; Gierk, B. [Development of a 12-item version of the OPD-Structure Questionnaire (OPD-SQS)]. Z Psychosom. Med. Psychother. 2015, 61, 262–274.

Ehrenthal, J.C.; Kruse, J.; Schmalbach, B.; Dinger, U.; Werner, S.; Schauenburg, H.; Brähler, E.; Kampling, H. Measuring personality functioning with the 12-item version of the OPD-Structure Questionnaire (OPD-SQS): Reliability, factor structure, validity, and measurement invariance in the general population. Front. Psychol. 2023, 14, 1248992.

Ehrenthal, J. C., Dinger, U., Lamla, A., Funken, B., & Schauenburg, H. (2008). Evaluation of the German version of the attachment questionnaire" Experiences in Close Relationships--Revised"(ECR-RD). Psychotherapie, psychosomatik, medizinische psychologie, 59(6), 215–223.

Fonagy P, Luyten P, Allison E. Epistemic petrification and the restoration of epistemic trust: A new conceptualization of borderline personality disorder and its psychosocial treatment. J Personal Disord. 2015;29(5):575–609. pmid:26393477

Fonagy, P., Campbell, C., Constantinou, M., Higgitt, A., Allison, E., & Luyten, P. (2022). Culture and psychopathology: An attempt at reconsidering the role of social learning. Development and Psychopathology, 34(4), 1205–1220.

Fricker M. Epistemic injustice: Power and the ethics of knowing. Oxford University Press; 2007.

Giddens A. The Consequences of Modernity. Stanford: Stanford University Press; 1990. 188 p.

Greiner, Christian, et al. "Epistemic Trust, Mistrust and Credulity Questionnaire (ETMCQ) validation in French language: Investigating association with loneliness." medRxiv (2024): 2024–05.

Henrich, J. (2020). The Weirdest People in the World: How the West Became Psychologically Peculiar and Particularly Prosperous. Allen Lane, Penguin Books

Hinz, A.; Klein, A.M.; Brähler, E.; Glaesmer, H.; Luck, T.; Riedel-Heller, S.G.; Wirkner, K.; Hilbert, A. Psychometric evaluation of the Generalized Anxiety Disorder Screener GAD-7, based on a large German general population sample. J. Affect. Disord. 2017, 210, 338–344.

Kampling, H., Kruse, J., Lampe, A., Nolte, T., Hettich, N., Brähler, E., … & Riedl, D. (2022). Epistemic trust and personality functioning mediate the association between adverse childhood experiences and posttraumatic stress disorder and complex posttraumatic stress disorder in adulthood. Frontiers in Psychiatry, 13, 919191.

Kerber, A., Ehrenthal, J. C., Zimmermann, J., Remmers, C., Nolte, T., Wendt, L. P., … & Knaevelsrud, C. (2024). Examining the role of personality functioning in a hierarchical taxonomy of psychopathology using two years of ambulatory assessed data. Translational Psychiatry, 14(1), 340.

Kliem, Sören, Cedric Sachser, Anna Lohmann, Dirk Baier, Elmar Braehler, Harald Gündel, and Jörg M. Fegert. "Psychometric evaluation and community norms of the PHQ-9, based on a representative German sample." Frontiers in Psychiatry 15 (2024): 1483782.

Kocalevent, R.-D.; Hinz, A.; Brähler, E. Standardization of the depression screener Patient Health Questionnaire (PHQ-9) in the general population. Gen. Hosp. Psychiatry 2013, 35, 551–555.

Liotti, Marianna, et al. "Unpacking trust: the Italian validation of the epistemic trust, mistrust, and credulity questionnaire (ETMCQ)." Plos one 18.1 (2023): e0280328.

Löwe, Bernd, Oliver Decker, Stefanie Müller, Elmar Brähler, Dieter Schellberg, Wolfgang Herzog, and Philipp Yorck Herzberg. "Validation and standardization of the Generalized Anxiety Disorder Screener (GAD-7) in the general population." Medical care 46, no. 3 (2008): 266–274.

Origgi G. Epistemic Injustice and Epistemic Trust. Social Epistemology. 2012 Apr 1;26(2):221–35.

Fonagy P, Luyten P, Allison E. Epistemic Petrification and the Restoration of Epistemic Trust: A New Conceptualization of Borderline Personality Disorder and Its Psychosocial Treatment. J Pers Disord. 2015 Oct;29(5):575–609. pmid:26393477

Löwe, B.; Spitzer, R.L.; Zipfel, S.; Herzog, W. Health-Questionnaire for Patients (PHQ-D). Complete Version and Short Form—Manual; Pfizer: Karlsruhe, Germany, 2002; Volume 2.

Luyten, P., Campbell, C., & Fonagy, P. (2020). Borderline personality disorder, complex trauma, and problems with self and identity: A social-communicative approach. Journal of Personality, 88(1), 88–105. 10.1111/jopy.12483

Luyten P, Campbell C, Allison E, Fonagy P. The Mentalizing Approach to Psychopathology: State of the Art and Future Directions. Annu Rev Clin Psychol. 2020 May 7;16(1):297–325. pmid:32023093

Mau, S., Lux, T., & Heide, J. (2024). Ost-und Westdeutsche für immer? Zu Wahrnehmungen von Unterschieden und Konflikten zwischen Ost-und Westdeutschen. KZfSS Kölner Zeitschrift für Soziologie und Sozialpsychologie, 76(1), 1–23.

Nolte, T., Hutsebaut, J., Sharp, C., Campbell, C., Fonagy, P., & Bateman, A. (2023). The role of epistemic trust in mentalization-based treatment of borderline psychopathology. Journal Of Personality Disorders.

R Core Team (2022). R: A language and environment for statistical computing. R Foundation for Statistical Computing, Vienna, Austria. URL https://www.R-project.org/

Pilkonis, P. A., Kim, Y., & Proietti, M. (1994). Adult Attachment Prototype Rating (AAPR). Unpublished manuscript. University of Pittsburgh.

Pilkonis, P. A., Kim, Y., Yu, L., & Morse, J. Q. (2014). Adult Attachment Ratings (AAR): An Item Response Theory Analysis. Journal of Personality Assessment, 96(4), 417–425. 10.1080/00223891.2013.832261

Preacher, K. J., & Hayes, A. F. (2008). Asymptotic and resampling strategies for assessing and comparing indirect effects in multiple mediator models. Behavior Research Methods, 40(3), 879–891. 10.3758/BRM.40.3.879

Preti, E., Richetin, J., Poggi, A., & Fertuck, E. (2023). A Model of Trust Processes in Borderline Personality Disorder: A Systematic Review. Current Psychiatry Reports, 25(11), 555.

Riedl, D., Rothmund, M., Grote, V., Fischer, M., Kampling, H., Kruse, J., … & Lampe, A. (2023). Mentalizing and epistemic trust as critical success factors in psychosomatic rehabilitation: results of a single center longitudinal observational study. Frontiers in Psychiatry, 14.

Riedl, D., Kampling, H., Nolte, T., Kirchhoff, C., Kruse, J., Sachser, C., … & Lampe, A. (2024). Utilization of Mental Health Provision, Epistemic Stance and Comorbid Psychopathology of Individuals with Complex Post-Traumatic Stress Disorders (CPTSD)—Results from a Representative German Observational Study. Journal of Clinical Medicine, 13(10), 2735.

Smits, M. L., De Vos, J., Rüfenacht, E., Nijssens, L., Shaverin, L., Nolte, T., … & Bateman, A. (2024). Breaking the cycle with Mentalization-Based Treatment Trauma-Focused: theory and practice of a trauma-focused group intervention. Frontiers in Psychology.

Sperber D, Clément F, Heintz C, Mascaro O, Mercier H, Origgi G, et al. Epistemic vigilance. Mind & language. 2010;25(4):359–93.

Spitzer, R.L.; Kroenke, K.; Williams, J.B.; Löwe, B. A brief measure for assessing generalized anxiety disorder: The GAD-7. Arch. Intern. Med. 2006, 166, 1092–1097.

Tabachnick, B.G., & Fidell, L.S. (2012). Using multivariate statistics. Boston: Pearson.

Simpson, J. A., & Rholes, W. S. (2002). Adult attachment, stress, and romantic relationships. Journal of Personality and Social Psychology, 83(3), 637–653. 10.1037/0022-3514.83.3.637

Strauß, B., & Lobo-Drost, A. (1999). Erwachsenen-Bindungsprototypen-Rating (EBPR): Eine Methode zur Erfassung von Bindungsqualitäten im Erwachsenenalter basierend auf dem Adult Attachment Prototype Rating von Pilkonis. Unter Mitarbeit von Elke Daudert, Diether Höger, Silke Schmidt & Jochen Eckert. Unveröffentlichtes Manual, Version 1.2, Jena/Hamburg, Januar 1999.

Tironi, Marta, et al. "Adverse childhood experiences and psychological maladjustment in adolescence: The protective role of epistemic trust, mentalized affectivity, and reflective functioning." Journal of Clinical Psychology (2024).

Warttig, S. L., Forshaw, M. J., South, J., & White, A. K. (2013). New, normative, English-sample data for the short form perceived stress scale (PSS-4). Journal of health psychology, 18(12), 1617-1628.

Weiland, A. M., Taubner, S., Zettl, M., Bartmann, L. C., Frohn, N., Luginsland, M., & Volkert, J. (2024). Epistemic trust and associations with psychopathology: Validation of the German version of the Epistemic Trust, Mistrust and Credulity-Questionnaire (ETMCQ). PloS one, 19(11), e0312995.

Wendt, L. P., Jankowsky, K., Schroeders, U., Nolte, T., Fonagy, P., Montague, P. R., … & Olaru, G. (2022). Mapping established psychopathology scales onto the Hierarchical Taxonomy of Psychopathology (HiTOP). Personality and Mental Health.

Wingenfeld, K.; Schäfer, I.; Terfehr, K.; Grabski, H.; Driessen, M.; Grabe, H.; Löwe, B.; Spitzer, C. Reliable, valide und ökonomische Erfassung frulher Traumatisierung: Erste psychometrische Charakterisierung der deutschen Version des Adverse Childhood Experiences Questionnaire (ACE). Psychother. Psychosom. Med. Psychol. 2011, 61, e10–e14.

Witt, A., Sachser, C., Plener, P. L., Brähler, E., & Fegert, J. M. (2019). The prevalence and consequences of adverse childhood experiences in the German population. Deutsches Ärzteblatt International, 116(38), 635.

Zeegers M, Colonnesi C, Stams G, Meins E. Mind matters: A meta-analysis on parental mentalization and sensitivity as predictors of infant-parent attachment. Psychological Bulletin. 2017; 143(12): 1245–72. pmid:28805399

